# Estimation of Rotavirus Vaccine Effectiveness Based on Whole Genome Sequences

**DOI:** 10.1101/2024.10.03.24314836

**Authors:** Jiye Kwon, Jose Jaimes, Mary E. Wikswo, Eileen J. Klein, Mary Allen Staat, James D. Chappell, Geoffrey A. Weinberg, Christopher J Harrison, Rangaraj Selvarangan, Coreen Johnson, Daniel M. Weinberger, Joshua L. Warren, Mathew D. Esona, Michael D. Bowen, Virginia E. Pitzer

## Abstract

**Introduction:** Rotavirus vaccine evaluations have noted small differences in vaccine effectiveness (VE) against rotavirus genotypes, defined by the two outer capsid proteins (VP7 or G-type and VP4 or P-type). However, the genomic landscape of group A rotavirus (RVA) and the impact of the remaining nine genome segments (i.e., the “backbone”) on VE are not fully understood. We incorporated whole genome sequence (WGS) data to characterize viruses responsible for rotavirus-associated gastroenteritis (RVGE) between vaccinated and unvaccinated individuals in the United States (U.S.).

**Methods:** We analyzed 254 RVGE cases with WGS data from seven U.S. New Vaccine Surveillance Network sites during 2012-2016. Using a “sieve analysis” framework, we evaluated the variability in vaccine protection based on genetic distance (GD) defined at WGS-level as the percent nucleotide difference between each case strain and the vaccine strain(s). Strain-specific VE estimates were calculated using the test-negative design, controlling for potential cofounders. Separate analyses were performed for the monovalent Rotarix® vaccine (RV1, GlaxoSmithKline) and the pentavalent RotaTeq® vaccine (RV5, Merck & Co.). We also examined the site-specific genetic diversity of circulating RVA strains in relation to vaccine coverage.

**Results:** RV1-vaccinated cases were more likely to be infected with strains with greater than 9.6% GD from the RV1 vaccine strain (unadjusted OR = 3.03, 95% confidence interval (CI): 1.15, 8.03). Strains with a genogroup 1 (Wa-like) backbone represented the majority (99%) of cases below the threshold, whereas more distant strains had genetic backbones that resembled the genogroup 2 (DS-1-like) and reassortant strains. The RV1 vaccine showed evidence of substantially better protection against strains with lower GD to the RV1 strain (VE = 80%, 95% CI: 68%, 89%) compared to more distant strains (VE = 51%, 95% CI: = -29%, 82%). RV5 demonstrated a similar but less pronounced pattern of better protection against strains with a lower minimum GD to the vaccine strains. Sites with higher RV1 usage showed a shift in strain distribution towards greater GD from the RV1 strain, with a similar trend observed for RV5.

**Conclusion:** Incorporating the complete genomic structure of RVA reveals that vaccine protection correlates with the diversity of non-outer capsid proteins. Our WGS-based analysis more clearly differentiated vaccine protection than analyses based on VP7 and VP4 alone. With more RVA vaccines in the pipeline, understanding the contribution of all gene segments to immune protection will be key to ensuring the long-term success of RVA vaccination programs.

## Introduction

Group A rotaviruses (RVA) remain the leading cause of death worldwide due to diarrheal disease in children under five years of age despite the availability of vaccines^1^. The virus causes acute RVA-associated gastroenteritis (RVGE), marked by severe dehydrating diarrhea. In high-income countries such as the United States (U.S.), the introduction of RVA vaccines has led to a substantial reduction in RVGE-associated morbidity, averting over 40,000 hospitalizations annually^2^. However, RVGE-associated morbidity remains a substantial concern both in the U.S. and globally^3^. RVA infection is imperfectly immunizing^4^, and in the U.S., RVA activity persists in a biennial pattern post-vaccine introduction.^5^ Globally, despite the high vaccine effectiveness (VE) observed in countries with low overall child mortality rates, far less optimal vaccine performance has been recorded in countries with higher overall child mortality rates^6,7^.

RVA is a double-stranded RNA virus, with its genome comprised of 11 segments, each encoding a viral protein^8^. All eleven genes represent the whole genome classification system of RVA genotype constellations as Gx-P[x]-Ix-Rx-Cx-Mx-Ax-Nx-Tx-Ex-Hx^9,10^. However, only the two outer capsid proteins (VP7 or G-type and VP4 or P-type) have been used historically to define RVA genotypes. RVA strains infecting humans have been characterized with a variety of G-type and P-type combinations^11^. While more than 36 G and 50 P types have been identified, G- and P-types G1, G2, G3, G4, G9, G12, P[4], P[6], and P[8] characterize the majority of the human RVA strains^3^. In 2008, the traditional binomial classification system for RVA was changed to an 11-segment system with the recommendation from the Rotavirus Classification Working Group^10^. The differences in the genetic backbone (i.e., nine internal gene segments) characterize the two major genogroups of human RVA strains: genogroup 1 (Wa-like; Gx-P[x]- I1-R1-C1-M1-A1-N1-T1-E1-H1) and genogroup 2 (DS-1-like; Gx-P[x]-I2-R2-C2-M2-A2-N2-T2-E2-H2)^11,12^. In the U.S., genogroup 2 strains are typically associated with the G2P[4] genotype, while G1P[8], G3P[8], G9[8], G12P[8] and other P[8] types are typically found on genogroup 1 strains.

Two live attenuated, oral rotavirus vaccines are currently licensed and recommended by the Advisory Committee on Immunization Practices (ACIP) in the U.S.^13^. RotaTeq^®^ (RV5; Merck) is a three-dose pentavalent bovine-human reassortant vaccine^14^. RV5 contains five strains that express outer capsid proteins commonly known to circulate in humans - G1, G2, G3, G4, P[8]; five to six of the nine-segment backbone of RV5 strains are shared with genogroup 2 strains^11^. Rotarix^®^ (RV1; GlaxoSmithKline) is a two-dose monovalent vaccine based on a live-attenuated human-derived G1P[8] strain with genogroup 1 backbone. Studies have identified small differences in VE against different G and P genotype combinations^15,16^. However, significant gaps remain in understanding the genomic landscape of RVA and the impact of the remaining nine genome segments (i.e., the “backbone”) on VE.

Prospective monitoring of VE is a crucial aspect of vaccination programs to assess impact and ensure continued effectiveness^17^. In the U.S., the New Vaccine Surveillance Network (NVSN) has been conducting pre- and post-vaccination surveillance for RVGE beginning in 2006^18^. As part of the active surveillance, NVSN also generates genomic information on case strains, including genotyping and nucleotide-level whole genome sequencing^3,19^. Genomic data offer high-resolution insight into the virus’ evolution, selection, and diversification^20,21^. These sequencing efforts provide an opportunity to better understand the factors and sequence-level biomarkers contributing to vaccine protection^20^.

Following vaccine introduction, the genomic analysis of RVA strains at both genotype and whole-genome levels provided valuable insights into the evolutionary patterns of circulating strains and revealed targets in the sequences that may potentially affect vaccine performance, thus requiring ongoing surveillance^3,22^. In this study, we take a further step by applying statistical models to directly estimate differences in vaccine protection based on whole genome sequence (WGS) data that encompass all eleven genes. We had three key objectives: 1) evaluate whether RVA vaccines offer differential protection against various RVA strains at the whole-genome level; 2) examine how site-level differences in RVA vaccine use correlate with RVA genetic diversity; and 3) identify factors and linkages at the whole-genome level that may predict VE.

## Methods

### Data sources and genetic sequences

We used epidemiological and WGS data collected from seven NVSN sites in the U.S. during December 2012 to June 2016. The seven NVSN sites include Seattle, Rochester, Nashville, Houston, Cincinnati, Kansas City, and Oakland. Comprehensive details on the NVSN acute gastroenteritis (AGE) surveillance methodology are available elsewhere^18,23^. In brief, following informed consent from a parent or guardian, the NVSN surveillance sites enrolled children 15 days to under the age of 18 who were admitted to the hospital, visited the emergency department (ED), or outpatient clinic with AGE (defined as ≥3 episodes of diarrhea and/or ≥1 episode of vomiting within 24 hours) within 10 days of onset. Demographic and clinical data, including vaccination status, were collected for each child. RVA infection status was determined using Premiere Rotaclone enzyme immunoassay (EIA) (Meridian Bioscience) in each site.

Positive strains were then sent to the Centers for Disease Control and Prevention (CDC) for genotype determination and whole genome sequencing^19^.

The whole-genome analysis and bioinformatics pipeline employed for the NVSN samples have been described previously^3^. Vaccine strain sequences, including one sequence for the monovalent RV1 vaccine ^24,25^ and five sequences for the pentavalent reassortant RV5 vaccine ^26^, were obtained from GenBank.

### Epidemiological data

Cases were defined as individuals with AGE who tested positive for RVA infection by EIA and who had samples sequenced at the WGS level. Controls were selected based on the test-negative design (TND) definition^17^ as individuals experiencing AGE who met all criteria for NVSN AGE surveillance and had samples collected, but who tested negative for RVA infection by EIA^27^. We excluded RVA-positive cases for whom WGS information was not available.

Vaccination status was obtained from regional immunization information systems and/or healthcare providers’ records as appropriate. Unvaccinated individuals and individuals who received at least one dose of either RV1 or RV5 were included in this study. For both cases and controls, we excluded those who received mixed doses (both RV1 and RV5) (n = 3 cases; 331 controls) or had unknown vaccine type (n = 22 cases; 770 controls).

For quality control purposes, cases with co-infections involving more than one RVA strain were excluded (n = 2). Additionally, cases that were suspected of shedding vaccine-associated strains were operationally defined as those exhibiting less than 2% overall nucleotide difference against the vaccine strains^28^ and were excluded from the analysis (n = 7).

### Vaccine protection estimation

Vaccine protection against RVA was estimated in two ways. First, we utilized a sieve analysis framework to assess relative VE against different RVA strains, relying solely on case information to compare WGS data for strains infecting unvaccinated and vaccinated individuals. Second, we incorporated information on the prevalence of vaccination from test-negative controls to estimate strain-specific VE for each of the genetic distance (GD)-based strain categories. GD was defined using p-distance (nucleotide) where we calculate the proportion of nucleotide sites different between the case strain and the vaccine strain.

The primary objective of the sieve analysis is to understand variability in vaccine protection across different strains^29^. Sieve analysis is a flexible statistical method that was initially devised to identify the differences in immune responses to vaccines by comparing the viral sequences between vaccinated and unvaccinated individuals in clinical trials^29,30^. The framework assesses whether vaccine protection against clinically significant infections depends on pathogen characteristics^31^, in this case defined by genomic sequences. This can help to identify potential sequence-based biomarkers that are informative of reduced vaccine protection. Notably, its flexibility allows the customization of strain categories without predefined bins^30^, accommodating variations beyond binomial classification based on the two outer capsid proteins (GxPx).

At the WGS level, we evaluated vaccine protection based on the GD between the case strain and the vaccine strain defined by percent nucleotide differences. Primary analysis focused on nucleotide-level analysis, since previous studies have shown that amino acid substitutions in RVA may be under strong purifying selection especially in VP4 and VP7 segments^3^. We created strain categories based on this nucleotide GD, conducting separate sieve analyses for the RV1 and RV5 vaccines. For the RV1 analysis, GD was calculated between the case strain and the monovalent G1P[8] strain, while for RV5, we used the minimum GD between the case strain and any of the five strains included in the pentavalent vaccine.

Strains were categorized based on k-means clustering^32^ across the distribution of GD. For each identified cluster (Strain_j_, where j=0 denotes the cluster closest to the vaccine strain and j=1, 2, 3, etc. characterize increasingly distant clusters identified through the k-means clustering), the maximum distance of the cluster was used as a cut-off (i.e., threshold) for the next strain category. Finally, the sieve analysis framework was implemented using a multinomial logistic regression (MLR) model^29^ to estimate the effect of vaccination status on the relative odds of infection with strain_j =1,2,3…_ versus strain_j=0_. We hypothesized that vaccine protection is more robust against closely related strains. We repeated the analysis for each of the 11 RVA genome segments in addition to the WGS.

We then estimated VE by comparing the prevalence of vaccination among RVA-positive RVGE cases and test-negative AGE controls. We used a logistic regression model to calculate the adjusted odds ratio of vaccination for cases versus controls and its 95% confidence interval (CI) adjusting for age (in years), sample collection year, and clinical setting (i.e., outpatient, inpatient, ED)^17,27^. Individual-level vaccination status was determined at the time of stool collection. An individual was classified as vaccinated if they had received at least one dose of RVA vaccine at the time of stool collection (and >14 days before the onset of AGE episode) and unvaccinated if they had not received any RVA vaccine or received their first dose <14 days before the onset of AGE.

The analysis was performed across three levels for the vaccination status of cases and controls – an overall analysis in which individuals were considered vaccinated if they received at least one dose of either RV1 or RV5; an RV1-specific analysis which included only RV1- vaccinated and unvaccinated individuals; and an RV5-specific analysis which included only

RV5-vaccinated and unvaccinated individuals. We estimated VE as one minus the adjusted odds ratio (i.e., the exponentiated coefficient of vaccination in the logistic regression model^33^).

For each of the three analyses described above (defined by vaccination status), we estimated the VE for all RVA cases and estimated strain-specific VE for the GD-based strain categories identified in the case-only sieve analysis. VE_any_ represents VE against any RVA strain; VE_j=0_ represents VE against strains genetically closer to the vaccine strain, and VE_j=1_ represents VE against strains more genetically distant from the vaccine strain.

### Sensitivity analyses

As a sensitivity analysis, we repeated our sieve analysis using amino acid-level distances for all segments. Additionally, for RV5-specific analyses, we expanded our approach to include average distance calculations to all five strains included in the vaccine rather than minimum distance to any of the five strains.

### Site-specific differences in genetic diversity

Finally, we examined how differential vaccine protection could influence genetic diversity at the population level by examining the diversity in circulating RVA strains across the seven NVSN sites in relation to vaccine coverage. We calculated and compared the GD distribution of strains to the RV1 and RV5 vaccines across each of the seven sites. Vaccine utilization across the sites was determined by the proportion of test-negative controls who received at least one dose of RV1 or RV5 or who were unvaccinated.

## Results

### Characteristics of cases with WGS information and controls

Our study included a total of 254 RVGE cases with WGS information and 12,733 RVA- negative AGE controls (Table 1). Among the cases with WGS information, 30 were RV1- vaccinated, 119 were RV5-vaccinated, and 105 were unvaccinated. The characteristics of the cases are broken down by vaccination status (Table 1). The median [range] age of RVA cases was 26 months [range: 0 - 174]. Across seven collection sites over the five years included in our study from 2012 - 2016, samples with WGS information were collected most from Nashville (54%) and least from Oakland (4%). The majority of samples were collected in 2013 (40%), and 64% of the sequenced samples were from ED patients. G12P[8] was the most common genotype (63%) among the sequenced cases, followed by G1P[8] (19%); the least collected genotypes were G2P[8] and G1P[4] (<1%).

**Table 1.**
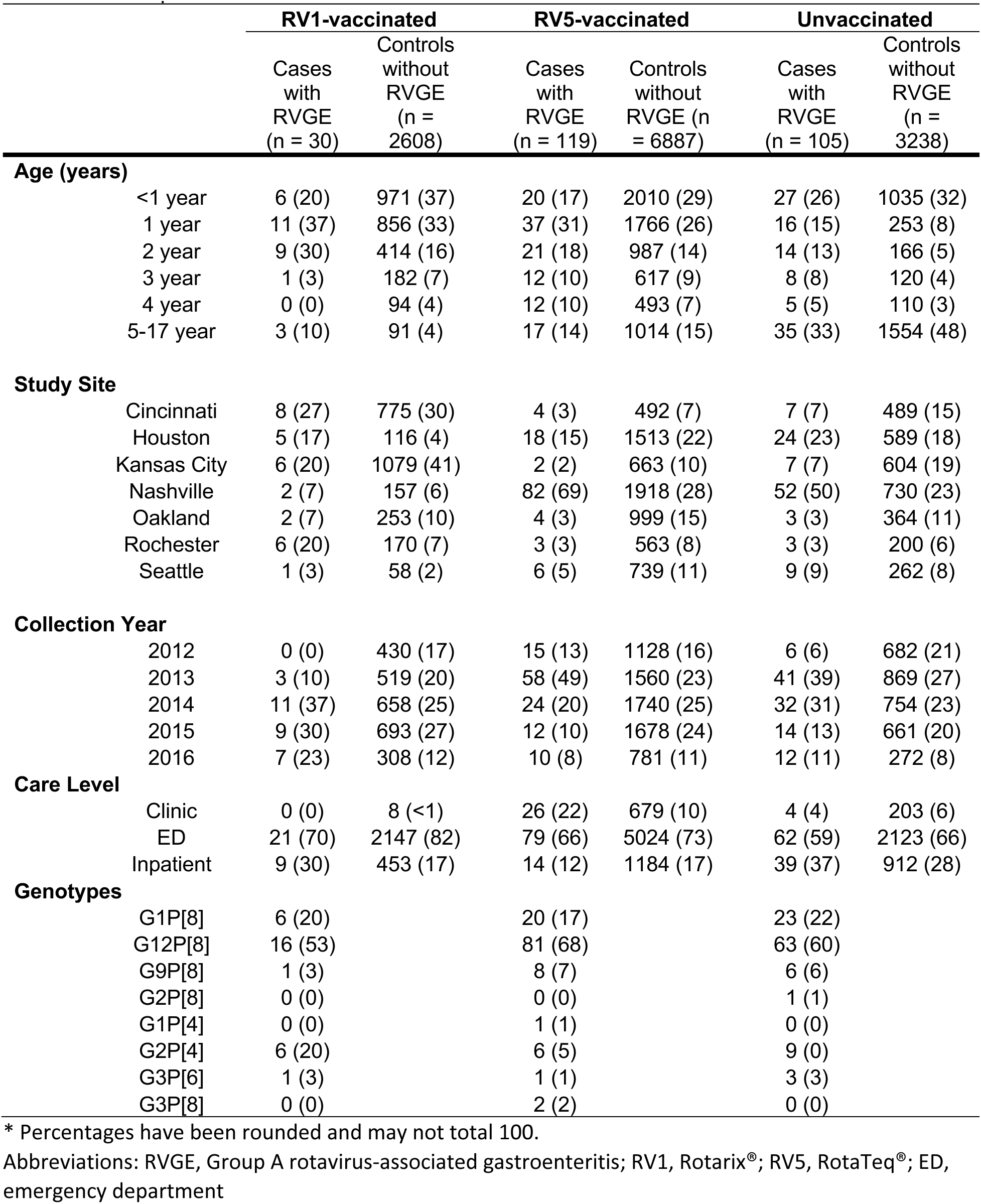
Description of New Vaccine Surveillance Network Rotavirus Cases and Controls*.

|  | RV1-vaccinated |  | RV5-vaccinated |  | Unvaccinated |  |
| --- | --- | --- | --- | --- | --- | --- |
|  | Cases with RVGE (n = 30) | Controls without RVGE (n = 2608) | Cases with RVGE (n = 119) | Controls without RVGE (n = 6887) | Cases with RVGE (n = 105) | Controls without RVGE (n = 3238) |
| **Age (years)** |  |  |  |  |  |  |
| <1 year | 6 (20) | 971 (37) | 20 (17) | 2010 (29) | 27 (26) | 1035 (32) |
| 1 year | 11 (37) | 856 (33) | 37 (31) | 1766 (26) | 16 (15) | 253 (8) |
| 2 year | 9 (30) | 414 (16) | 21 (18) | 987 (14) | 14 (13) | 166 (5) |
| 3 year | 1 (3) | 182 (7) | 12 (10) | 617 (9) | 8 (8) | 120 (4) |
| 4 year | 0 (0) | 94 (4) | 12 (10) | 493 (7) | 5 (5) | 110 (3) |
| 5-17 year | 3 (10) | 91 (4) | 17 (14) | 1014 (15) | 35 (33) | 1554 (48) |
| **Study Site** |  |  |  |  |  |  |
| Cincinnati | 8 (27) | 775 (30) | 4 (3) | 492 (7) | 7 (7) | 489 (15) |
| Houston | 5 (17) | 116 (4) | 18 (15) | 1513 (22) | 24 (23) | 589 (18) |
| Kansas City | 6 (20) | 1079 (41) | 2 (2) | 663 (10) | 7 (7) | 604 (19) |
| Nashville | 2 (7) | 157 (6) | 82 (69) | 1918 (28) | 52 (50) | 730 (23) |
| Oakland | 2 (7) | 253 (10) | 4 (3) | 999 (15) | 3 (3) | 364 (11) |
| Rochester | 6 (20) | 170 (7) | 3 (3) | 563 (8) | 3 (3) | 200 (6) |
| Seattle | 1 (3) | 58 (2) | 6 (5) | 739 (11) | 9 (9) | 262 (8) |
| **Collection Year** |  |  |  |  |  |  |
| 2012 | 0 (0) | 430 (17) | 15 (13) | 1128 (16) | 6 (6) | 682 (21) |
| 2013 | 3 (10) | 519 (20) | 58 (49) | 1560 (23) | 41 (39) | 869 (27) |
| 2014 | 11 (37) | 658 (25) | 24 (20) | 1740 (25) | 32 (31) | 754 (23) |
| 2015 | 9 (30) | 693 (27) | 12 (10) | 1678 (24) | 14 (13) | 661 (20) |
| 2016 | 7 (23) | 308 (12) | 10 (8) | 781 (11) | 12 (11) | 272 (8) |
| **Care Level** |  |  |  |  |  |  |
| Clinic | 0 (0) | 8 (<1) | 26 (22) | 679 (10) | 4 (4) | 203 (6) |
| ED | 21 (70) | 2147 (82) | 79 (66) | 5024 (73) | 62 (59) | 2123 (66) |
| Inpatient | 9 (30) | 453 (17) | 14 (12) | 1184 (17) | 39 (37) | 912 (28) |
| **Genotypes** |  |  |  |  |  |  |
| G1P[8] | 6 (20) |  | 20 (17) |  | 23 (22) |  |
| G12P[8] | 16 (53) |  | 81 (68) |  | 63 (60) |  |
| G9P[8] | 1 (3) |  | 8 (7) |  | 6 (6) |  |
| G2P[8] | 0 (0) |  | 0 (0) |  | 1 (1) |  |
| G1P[4] | 0 (0) |  | 1 (1) |  | 0 (0) |  |
| G2P[4] | 6 (20) |  | 6 (5) |  | 9 (9) |  |
| G3P[6] | 1 (3) |  | 1 (1) |  | 3 (3) |  |
| G3P[8] | 0 (0) |  | 2 (2) |  | 0 (0) |  |

\* Percentages have been rounded and may not total 100.
Abbreviations: RVGE, Group A rotavirus-associated gastroenteritis; RV1, Rotarix®; RV5, RotaTeq®; ED, emergency department

Among RVA-negative AGE controls, 2608 (20%), 6887 (54%), and 3238 (25%) were RV1-vaccinated, RV5-vaccinated, and unvaccinated, respectively. Overall, controls were younger than cases; 32% were from children <1 year old, followed by 1 year of age (23%) and older than 5 (21%). The site with the most RVA-negative controls was Nashville (22%), whereas Rochester (7%) and Seattle (8%) contributed the fewest controls. The majority of the control samples were from ED patients (73%).

### Genetic distance of case strains to RV1 and RV5 vaccine strains

For the 135 cases included in the RV1 analysis, minimum GD to RV1 vaccine strain ranged from 2.9% to 22.0% (Figure 1a). For the 224 cases included In the RV5 analysis, the minimum GD to any of the RV5 strains ranged from 14.9% to 18.3% (Figure 1b).

**Figure 1.**
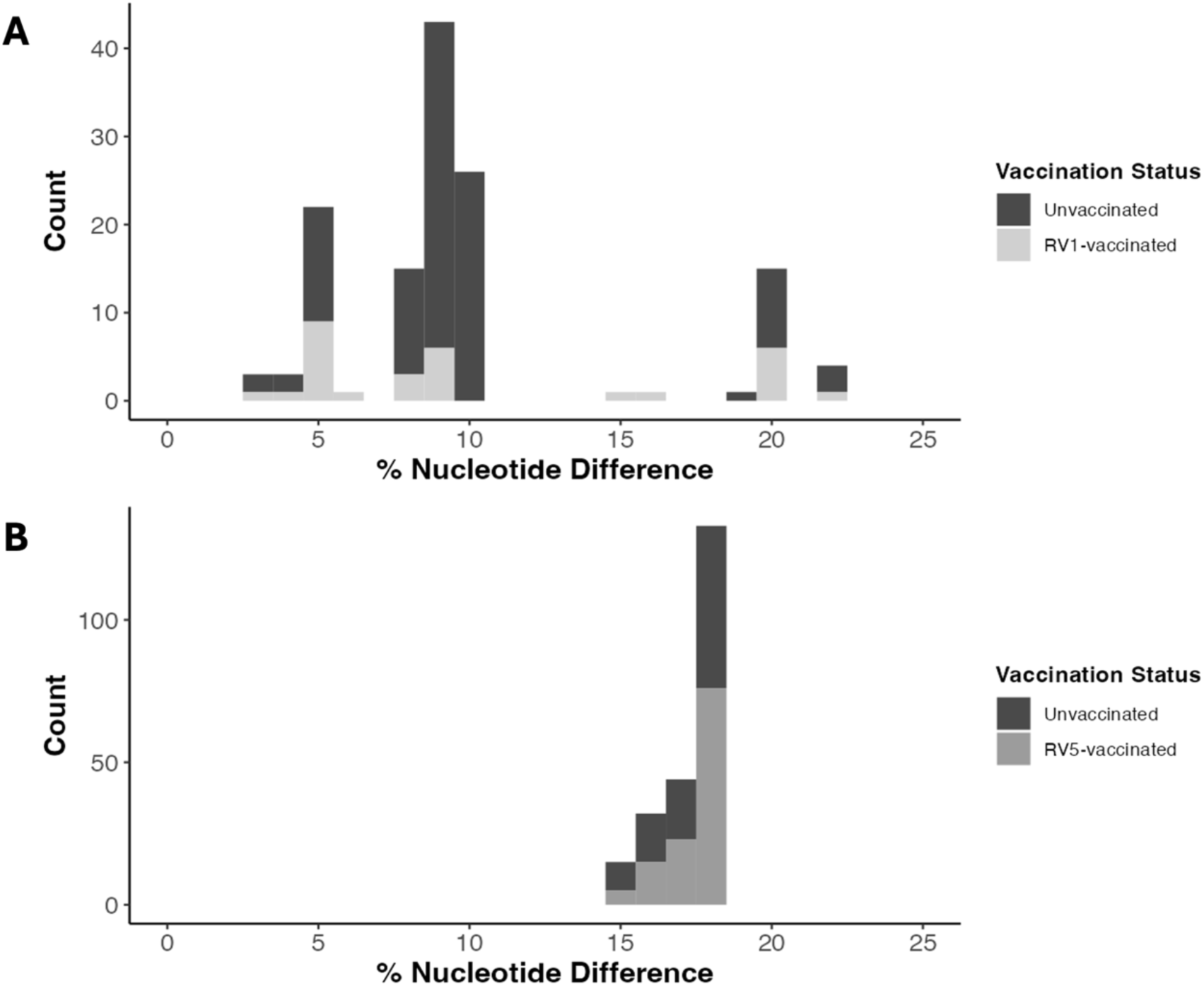
Nucleotide-level genetic distance (GD) distribution of all cases colored by vaccination status for the vaccine-specific analyses. (A) The RV1-specific analysis includes cases who received at least one dose of RV1 vaccine or were unvaccinated. GD is represented as the percent nucleotide difference between the case and RV1 vaccine strain. (B) The RV5-specific analysis includes cases who received at least one dose of RV5 vaccine or were unvaccinated. GD is represented as the minimum percent nucleotide difference between the case strain to any of the five strains included in the pentavalent RV5 vaccine.

The K-means clustering algorithm binned each of the GD distributions into two strain categories for both the RV1 and RV5 analysis (Supplemental Figure 1). The identified thresholds between the two clusters were set at 9.6% for the RV1 analysis and 16.7% for the RV5 analysis. Incorporating this information, we assigned case strains with GD greater than or equal to the defined threshold as genetically distant (*j*=1). Conversely, those with GD less than the threshold were classified as *j*=0, representing case strains genetically closer to the vaccine strain.

In the RV1-specific analysis, among those infected with strains in the more distant *j*=1 cluster, 41% of cases were RV1-vaccinated, whereas only 19% of those infected with strains from the *j*=0 cluster were RV1-vaccinated. Based on the sieve analysis, RV1-vaccinated individuals who experienced RVGE were 3.03 times (95% CI: 1.15, 8.03) more likely to be infected with more distant strains compared to strains more closely related to the RV1 vaccine (unadjusted OR). When GD was defined at the amino acid level, the observed effect, while in a similar direction, was less pronounced (OR [95% CI]: 2.57 [0.95, 6.97]) (Supplemental Figure 2).

Meanwhile, the RV5-specific analysis showed a more similar distribution between RV5- vaccinated and unvaccinated individuals in both strain categories and less genetic variation overall (OR [95% CI]: 1.60 [0.85, 3.02]). Among those infected with strains in the more distant *j*=1 cluster, 56% of cases were RV5-vaccinated, whereas 44% of those infected with strains from the *j*=0 cluster were RV5-vaccinated; this suggests the RV5 vaccine provided more uniform protection against clinically significant RVGE across the observed GD distribution. Results were similar when the analysis was conducted using average distance to all RV5 strains (OR [95% CI]: 1.96 [0.78, 4.94]).

Individual segment-level analysis similarly exhibited consistent trends. Although the identified threshold and magnitude of the effect of vaccination varied across each segment, a similar pattern of variable protection across the GD distributions for each segment was evident (Supplemental Figures 3 & 4; Supplemental Table 1). When comparing the two vaccine-specific analyses, the RV1-specific analysis revealed a more distinct potential for genetic linkage across segments, with identical ORs (i.e., grouping of segments from vaccinated versus unvaccinated individuals) for VP1-3 and NSP3-4. There were also two segments (VP6 and NSP1) for which the OR was less than 1, suggesting vaccinated individuals were more likely to be infected with strains closely related to the RV1 vaccine. None of the segment-specific ORs were significant in the RV5 analysis. Overall, the WGS-level comparison captured the relative vaccine protection across the full distribution of GD more effectively than the individual segment-level analysis.

### Genomic characterization of distance-based strain categories

For each vaccine-specific analysis, we examined the genomic composition of each GD-based strain at two levels – based on the outer capsid-protein genotype (GxPx) (Figure 2a, Supplemental Figure 6a) and the genogroup of the other nine backbone segments, i.e., genotype constellation^11^ (Figure 2b, Supplemental Figure 6b).

**Figure 2.**
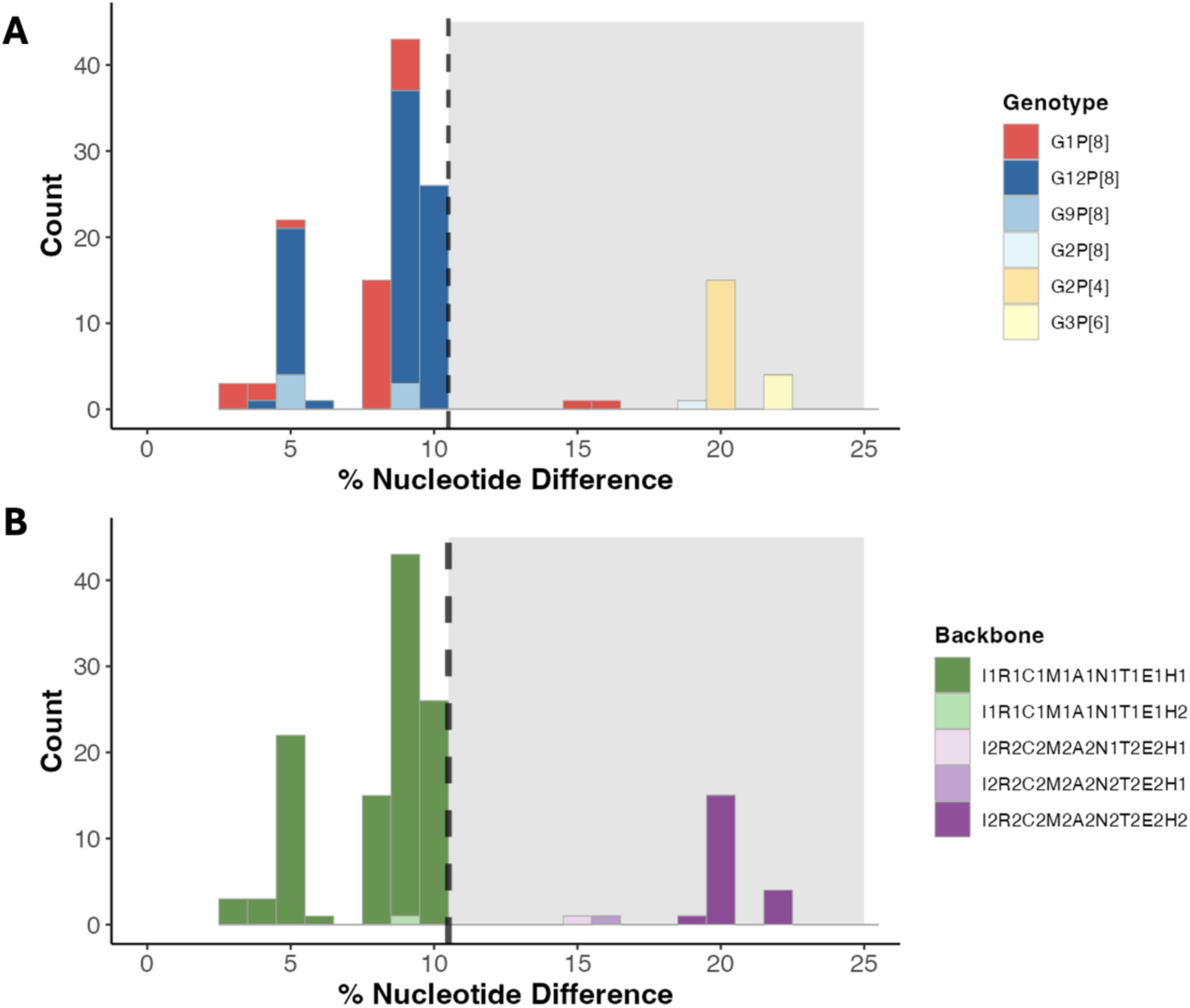
Characterization of the nucleotide-level genetic distance (GD) distribution for cases in the RV1-specific analysis according to genotype and genogroup. Strains are colored according to (A) GxPx genotype information, and (B) genogroup of the remaining nine-segment backbone. GD is represented as percent nucleotide difference between the case and RV1 vaccine strain. The black dotted line and grey background represent the threshold dividing Strain_j=0_ vs Strain_j=1._ Histogram binwidth is set to 1 and due to rounding issues, the threshold line in the figure is set at 10.5 for better visualization; for actual analysis, the threshold is set at 9.6%.

In RV1 analysis, the GD distribution stratified by genotype showed that strains below the 9.6% GD threshold (Strain_j=0_) were predominantly G12P[8] (70%), followed by G1P[8] (24%) and G9P[8] (6%) (Figure 2a). Strains above the 9.6% GD threshold (Strain_j=1_) were predominantly G2P[4] (68%) as well as a mix of G3P[6] (18%), G1P[8] (9%), and G2P[8] (5%). It is noteworthy that G1P[8] strains (homotypic to RV1) were observed on both sides of the threshold; moreover, the two reassortant G1P[8] strains above the GD threshold were from RV1-vaccinated individuals. A summary of GD for each RVA genotype is shown in Supplemental Table 2.

When the genogroup constellation of the nine backbone segments was considered, strains with a genogroup 1 backbone (I1-R1-C1-M1-A1-N1-T1-E1-H1) represented the majority (99%) of cases below the threshold (Strain_j=0_). Strains above the threshold (Strain_j=1_) had genetic backbones that resembled genogroup 2 (I2-R2-C2-M2-A2-N2-T2-E2-H2) and reassortant strains that were predominantly DS-1-like. The genogroup of the nine backbone segments (Fig. 2b) more clearly differentiated between the two strain categories compared to VP7 and VP4 (G- & P-type) only (Fig. 2a). Overall, when sensitivity analysis using percent amino acid differences was conducted, a consistent pattern was observed, with the exception of one G1P[8] strain that possessed a genogroup 1 and 2 reassortant backbone and was categorized as belonging to the Strain_j=0_ group (see Supplemental Figure 5).

In the RV5 analysis, the distinction between genotypes and the genogroup constellations of the backbone segments on either side of the threshold was less clear, although strains above the threshold were predominantly G12P[8] with a genogroup 1 backbone (Supplemental Figure 6). While sieve analysis revealed no notable differences between minimum and average distances to RV5 strains, visual patterns in genotype and genogroup were evident across the GD distribution. Notably, we observed distinctly opposite patterns compared to RV1-specific analysis when using the average distance to all RV5 strains, with strains above the threshold mostly coming from genogroup 1 (Supplemental Figure 7).

### Genetic distance of all case strains to the RV1 vaccine strain

When we considered the GD distribution of strains from all cases (including RV5- vaccinated cases, n = 254) to the RV1 vaccine strain, we found 30% of strains above the threshold (GD > 9.6%) were from RV1-vaccinated cases, 27% were from RV5-vaccinated cases, and 43% were from unvaccinated cases, whereas 9%, 50%, and 41% of strains below the threshold were from RV1-vaccinated, RV5-vaccinated, and unvaccinated cases, respectively (Supplemental Figure 8). Similar to the RV1 vaccine-specific analysis, the vast majority of strains below the threshold had a genogroup 1 backbone, whereas strains above the threshold had a genogroup 2 or reassortant backbone; again, G1P[8] strains were found on both sides of the threshold (Supplemental Figure 9). Overall, fully heterotypic strains, including G3P[6] and G2P[4], exhibited the furthest median distance from the G1P[8] RV1 vaccine strain (Supplemental Figure 10). Partially heterotypic strains were genetically closer to the vaccine strain than the fully heterotypic strains but displayed a wider distribution of distances, indicated by higher within-genotype standard deviation and broader range. Interestingly, homotypic G1P[8] strains demonstrated the widest range of GD to the RV1 vaccine strain, spanning both sides of the threshold.

### Distance-based strain-specific vaccine effectiveness estimates

RV1 demonstrated differential vaccine protection against strains above versus below the threshold in the GD distribution, whereas no such differential protection was observed in RV5 analysis. The overall VE of at least one dose of either RVA vaccine against any RVA strain (VE_any_) over the five-year study period was estimated to be 62% (95% CI: 49%, 72%). Similar protection against any RVA strain was observed in RV1- and RV5-specific analyses (Table 2; Table 3). However, when we categorized RVA strains according to GD to the RV1 vaccine strains, we found that the RV1 vaccine exhibited substantially stronger protection against strains below the threshold (VE_j=0_ [95% CI] = 80% [68%, 89%]) compared to strains above the threshold (VE_j=1_ [95% CI] = 51% [-29%, 82%]). For strain categories based on minimum GD to the RV5 vaccine strains, we again observed stronger vaccine protection against strains below the threshold (VE_j=0_ [95% CI] = 73% [51%, 86%]) compared to strains above the threshold (VE_j=1_ [95% CI] = 55% [36%, 68%]) for the RV5-specific analysis, but the difference was less pronounced (Table 3). The results can be compared to VE estimates using genotype classification based on the two outer capsid proteins (Supplemental Table 3).

**Table 2.** Study-wide and strain-specific vaccine effectiveness (VE) estimates against RVGE from study years 2012 – 2016, based on genetic distance to the RV1 strain. In the overall analysis, individuals were considered vaccinated if they received at least one dose of either RV1 or RV5. In the RV1-specific analysis, only RV1-vaccinated and unvaccinated individuals w ere included.

|  | **Adjusted\***<br>**$VE_{any}$**<br>(95% CI) | P-value | **Adjusted\***<br>**$VE_{j=0}$**<br>(95% CI) | P-value | **Adjusted\***<br>**$VE_{j=1}$**<br>(95% CI) | P-value |
| --- | --- | --- | --- | --- | --- | --- |
| **Overall analysis** | 62% (49, 72) | < 0.001 | 61% (47, 71) | < 0.001 | 70% (31, 86) | 0.01 |
| **RV1-specific analysis** | 76% (63, 85) | < 0.001 | 80% (68, 89) | < 0.001 | 51% (-29, 82) | 0.146 |

\* Adjusted for age, care level (ED, inpatient, outpatient), collection year

**Table 3.** Study-wide and strain-specific vaccine effectiveness (VE) estimates against RVGE from study years 2012 – 2016, based on minimum genetic distance to any of the RV5 vaccine strains. In the overall analysis, individuals were considered vaccinated if they received at least one dose of either RV1 or RV5. In the RV5-specific analysis, only RV5- vaccinated and unvaccinated individuals were included.

|  | **Adjusted\***<br>**VE<sub>any</sub>**<br>(95% CI) | P-value | **Adjusted\***<br>**VE<sub>j=0</sub>**<br>(95% CI) | P-value | **Adjusted\***<br>**VE<sub>j=1</sub>**<br>(95% CI) | P-value |
| --- | --- | --- | --- | --- | --- | --- |
| **Overall analysis** | 62% (49, 72) | < 0.001 | 64% (38, 79) | < 0.001 | 62% (46, 73) | <0.001 |
| **RV5-specific analysis** | 60% (46, 71) | < 0.001 | 73% (51, 86) | < 0.001 | 55% (36, 68) | <0.001 |

\* Adjusted for age, care level (ED, inpatient, outpatient), collection year

### Differences in the geographical distribution of RVA strains across seven U.S. NVSN sites

At the population level, diversity in circulating RVA strains across the seven NVSN sites differed with vaccine coverage. While coverage with either vaccine was similar across the seven study sites (72% to 79%) the proportion of RV1 versus RV5 vaccine usage varied (Supplemental Table 4). Interestingly, among study sites where RV1 vaccine usage was higher (i.e., over 40% of controls were vaccinated with RV1; Supplemental Table 4), cases tended to be infected with strains that were more genetically distant from the RV1 vaccine compared to sites with higher RV5 use (Figure 3a). Specifically, Kansas City and Cincinnati had higher RV1 use, and 33% and 79% of cases from these sites, respectively, were infected with strains more distant from the RV1 vaccine (Supplemental Table 5). Conversely, more than two-thirds of controls in Nashville, Seattle, and Houston were vaccinated with RV5, and >80% of case strains from these sites had less than 9.6% GD to the RV1 vaccine strain (Supplemental Table 5). We also observed a similar trend of median GD shifting further away from the vaccine strain with varying degrees of RV5 use (Figure 3b). In the sensitivity analysis of the RV5-specific study, where average distance rather than minimum distance to RV5 vaccine strains was used, we observed a similar but less gradual pattern of divergence from the RV5 vaccine strains (Supplemental Figure 11).

**Figure 3.**
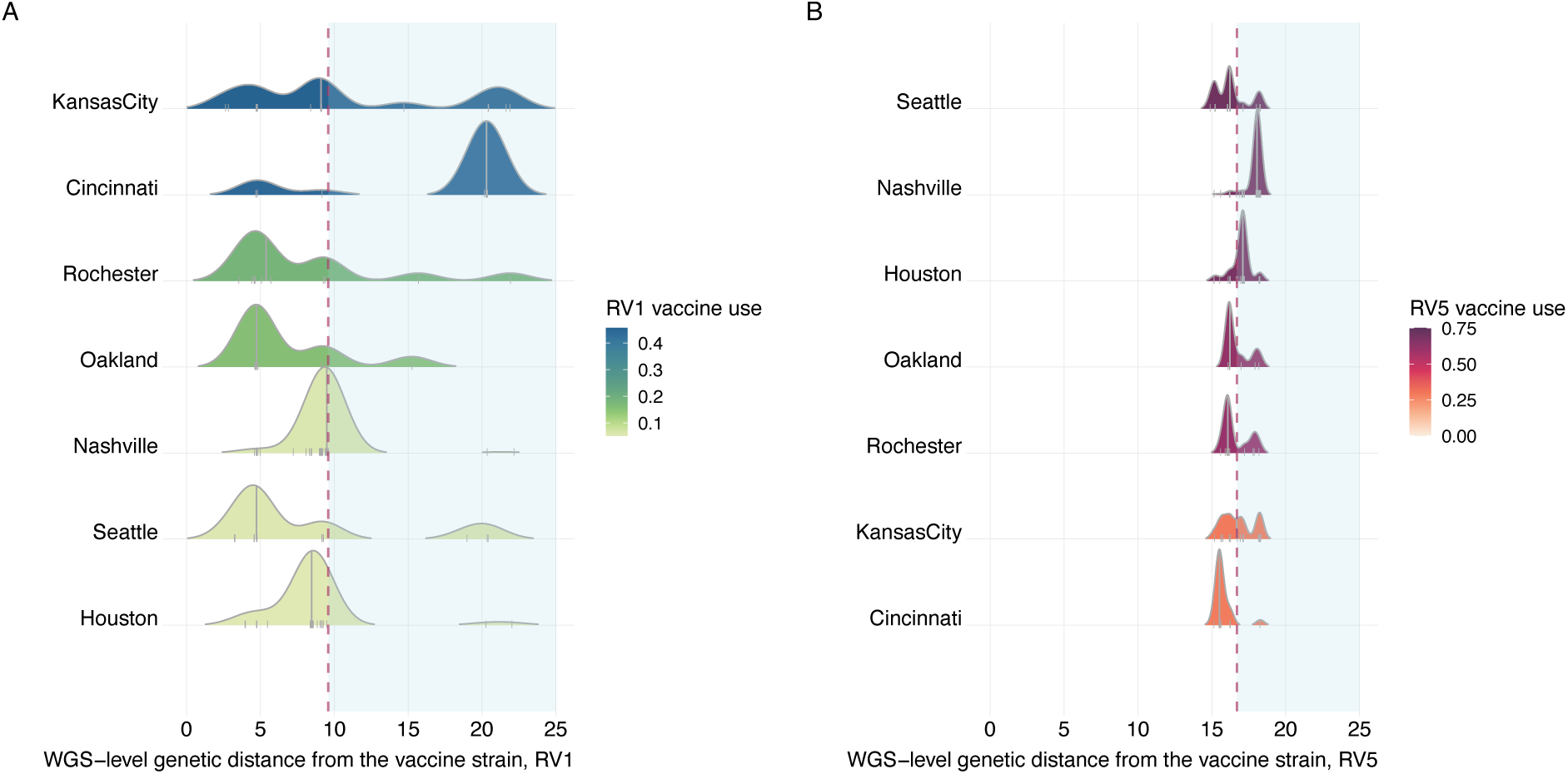
Nucleotide-level genetic distance distribution of all cases to the rotavirus vaccine strain* for each of the seven U.S. NVSN study sites (n= 254). *(A) RV1 vaccine and (B) minimum GD to any of the RV5 vaccine strains. Each site is colored by the proportion of vaccine usage amongst controls. The grey line in the distribution represents median GD for each study site. The red dotted line and light blue shaded area represents the threshold dividing Strain_j=0_ vs Strain_j=1._ Study sites are ordered by descending vaccine use (top to bottom).

## Discussion

Our analysis provides an improved understanding of how the full genomic composition of RVA contributes to vaccine protection. Leveraging a comprehensive dataset of WGSs collected across seven U.S. NVSN surveillance sites over five years, we calculated the GD between case strains and both the monovalent Rotarix® (RV1) and pentavalent RotaTeq® (RV5) vaccines widely used in the U.S. Our findings underscore that protection from RVA vaccines depends on genetic diversity beyond the outer capsid proteins. We highlight the importance of incorporating the entire genomic structure of RVA in understanding heterogeneity in vaccine protection.

Since the introduction of RVA vaccines in the U.S. in 2006, the impact of vaccine-induced immune pressure on the evolution of RVA in the post-vaccine era is still under close monitoring and active investigation^3^. Prior to this study, most VE analyses primarily focused on the two outer capsid proteins (VP7/G-type and VP4/P-type) to estimate genotype-specific VE against RVGE^34,35^. In our study, we evaluated all eleven segments of RVA regardless of pre- defined genotypes and translated the WGS information into a GD metric. Using this framework, we found that children vaccinated with RV1 were more likely to have breakthrough infections caused by strains more distant to the vaccine strain (i.e., higher GD). The GD threshold identified by k-means clustering clearly differentiated between strains with a predominantly genogroup 1 backbone (similar to the vaccine strain) versus those with a predominantly genogroup 2 backbone. Notably, RVGE cases infected with G1P[8] strains appeared on both sides of the GD threshold; further analysis revealed that two of these cases – both of whom were vaccinated – were infected with RV1-wild type reassortant strains that had a predominantly DS-1-like backbone. In contrast, there was no significant difference in the minimum GD of strains infecting RV5-vaccinated versus unvaccinated individuals.

When analyzing each segment, possible genetic linkages across segments was notable for the RV1 analysis, with two distinct clusters standing out. The larger cluster, which included segments VP1-3, NSP3, and NSP4, showed that vaccinated individuals were more likely to experience breakthrough infections caused by strains genetically distant from the vaccine strain. Conversely, the VP6 and NSP1 segments exhibited the opposite trend, which might be due to confounding by year of sample collection (which we control for in the main analysis) if both GD and vaccine coverage are increasing over time. Vaccinated individuals may also be more likely to have been infected with a vaccine-reassortant strain, e.g., for VP6, strains in the *j*=0 group show very small GD to the RV1 VP6 (Supplemental Figure 3). Previous studies have highlighted that VP6 is notably conserved, although recombination events have been observed^36,37^.

Additionally, VP6, VP3, and NSP1 segments have been reported to share overlapping tree space^38^. Nonetheless, translating the data into a GD metric at the WGS level made these patterns more apparent.

Nevertheless, our analysis suggests that in the post-licensure era of RVA vaccines, the combined use of RV1 and RV5 remains effective in preventing clinically significant RVGE. Comparing the vaccination status of test-negative controls to that of the RVGE cases with WGS data, we estimated the overall effectiveness of at least one dose of either RV1 or RV5 was 62% (95% CI: 49%,72%). Our estimates of VE were slightly lower than other studies utilizing NVSN data^15,27^. This discrepancy may stem from potential biases in the selection of samples to be sent for sequencing, potentially favoring cases from vaccinated individuals (e.g., to determine whether they were infected with vaccine-derived strains), leading to an overrepresentation of vaccinated cases and a bias in our estimates towards lower VE. Additionally, our definition of vaccinated individuals encompassed those who received at least one dose, whereas other studies only included individuals who completed a full course of RVA vaccines, potentially yielding better vaccine protection. Nevertheless, both our sieve analysis and adjusted VE estimates suggest that the RV1 vaccine may provide a more targeted strain-specific protection across the GD distribution. The RV1 vaccine showed evidence of substantially better protection against strains with a lower GD (Strain _j=0_) to the RV1 strain. The RV5 analysis also showed slightly better protection against strains with a smaller minimum GD to any RV5 strain, although the difference in VE was less pronounced.

Vaccination may exert selective pressures on viral populations^39,40^. Previous reports from several countries, including the U.S., Australia and Belgium, have highlighted changes in the RVA viral population following vaccination^41–43^. In particular, previous studies at the genotype level have indicated an increased proportion of G2P[4] strains following the introduction of the RV1 vaccine^44–46^. However, understanding whether these changes are directly influenced by vaccination and identifying what constitutes a “vaccine escape” strain remains a complex challenge. The Australian immunization program provided a unique natural experiment since different states elected to use RV1 versus RV5 during the initial rollout^44^. In the pre-vaccine era, G1P[8] was the dominant strain countrywide, while in the post-vaccine evaluation of RVA genotype diversity, differences in circulating strains between states using RV1 and RV5 were observed across the country^44^. We observed a similar pattern at the WGS level in the U.S. Study sites with a higher proportion of RV1 usage (such as Kansas City and Cincinnati) displayed a shift in the distribution of strains towards greater GD from the RV1 vaccine strain. In contrast, study sites with a higher proportion of RV5 use (e.g., Houston and Nashville) exhibited a distribution skewed towards strains more genetically similar to the RV1 vaccine strain and more distant from the RV5 strains. Our analysis provides further insights that RVA vaccines may exert selective pressure on the RVA population, especially when considering the WGS level.

Limitations to our study include the relatively small sample size of cases with WGS data, particularly evident in the RV1 analysis due to the limited use of RV1 compared to RV5 in the U.S. This is reflected in the wider confidence intervals within the RV1-specific analyses, particularly for the VE against strains above the GD threshold. To optimize our use of the limited sample size available, we adopted a broader case definition by not limiting our analysis to individuals who received a full course, which might have provided a conservative estimate of VE. Despite these limitations, our VE estimates remain consistent with other published studies. Future studies should seek to confirm our findings in other settings with WGS data available.

Our study introduces a robust framework for utilizing RVA WGS data to estimate variations in VE for RVA vaccines. It underscores the crucial significance of incorporating the full genomic structure, showcasing that vaccine protection correlates with genetic diversity beyond the outer capsid proteins. With four RVA vaccines currently available and more in the pipeline, this framework can aid in evaluating other RVA vaccines, enabling the identification of genomic components contributing to reduced vaccine protection. Decision-makers should consider the current genetic diversity of RVA to help guide vaccine selection and continue to monitor the genomic landscape as vaccines are rolled out.

## Acknowledgments

We would like to thank the following individuals for their helpful conversations and feedback on this work: Chrispin Chaguza, Nathan D. Grubaugh, Khuzwayo Jere, and Jelle Matthijnssens.

## Data availability statement

Nucleotide sequences of the complete genes from the study strains were retrieved from the publicly available GenBank database. The corresponding accession numbers can be found in the Supplemental File.

## References

1. Troeger C, Blacker BF, Khalil IA, et al. Estimates of the global, regional, and national morbidity, mortality, and aetiologies of diarrhoea in 195 countries: a systematic analysis for the Global Burden of Disease Study 2016. The Lancet Infectious Diseases. 2018;18(11):1211–1228. doi:10.1016/S1473-3099(18)30362-1

2. Tate JE, Panozzo CA, Payne DC, et al. Decline and change in seasonality of US rotavirus activity after the introduction of rotavirus vaccine. Pediatrics. Aug 2009;124(2):465–71. doi:10.1542/peds.2008-3528

3. Esona MD, Gautam R, Katz E, et al. Comparative genomic analysis of genogroup 1 and genogroup 2 rotaviruses circulating in seven US cities, 2014-2016. Virus Evol. Jan 2021;7(1):veab023. doi:10.1093/ve/veab023

4. Lopman BA, Pitzer VE. Waxing Understanding of Waning Immunity. J Infect Dis. Mar 5 2018;217(6):851–853. doi:10.1093/infdis/jix670

5. 5. Baker JM, Tate JE, Steiner CA, Haber MJ, Parashar UD, Lopman BA. Longer-term Direct and Indirect Effects of Infant Rotavirus Vaccination Across All Ages in the United States in 2000– 2013: Analysis of a Large Hospital Discharge Data Set. Clinical Infectious Diseases. 2018;68(6):976–983. doi:10.1093/cid/ciy580

6. Kirkwood CD, Ma LF, Carey ME, Steele AD. The rotavirus vaccine development pipeline. Vaccine. Nov 28 2019;37(50):7328–7335. doi:10.1016/j.vaccine.2017.03.076

7. Burnett E, Parashar UD, Tate JE. Real-world effectiveness of rotavirus vaccines, 2006-19: a literature review and meta-analysis. Lancet Glob Health. Sep 2020;8(9):e1195-e1202. doi:10.1016/s2214-109x(20)30262-x

8. Greenberg HB, Estes MK. Rotaviruses: from pathogenesis to vaccination. Gastroenterology. May 2009;136(6):1939–51. doi:10.1053/j.gastro.2009.02.076

9. Matthijnssens J, Ciarlet M, McDonald SM, et al. Uniformity of rotavirus strain nomenclature proposed by the Rotavirus Classification Working Group (RCWG). Arch Virol. Aug 2011;156(8):1397–413. doi:10.1007/s00705-011-1006-z

10. Matthijnssens J, Ciarlet M, Rahman M, et al. Recommendations for the classification of group A rotaviruses using all 11 genomic RNA segments. Arch Virol. 2008;153(8):1621–9. doi:10.1007/s00705-008-0155-1

11. Matthijnssens J, Van Ranst M. Genotype constellation and evolution of group A rotaviruses infecting humans. Curr Opin Virol. Aug 2012;2(4):426–33. doi:10.1016/j.coviro.2012.04.007

12. Matthijnssens J, Ciarlet M, Heiman E, et al. Full Genome-Based Classification of Rotaviruses Reveals a Common Origin between Human Wa-Like and Porcine Rotavirus Strains and Human DS-1-Like and Bovine Rotavirus Strains. Journal of Virology. 2008;82(7):3204–3219. doi:doi:10.1128/jvi.02257-07

13. Parashar UD, Alexander JP, Glass RI. Prevention of rotavirus gastroenteritis among infants and children. Recommendations of the Advisory Committee on Immunization Practices (ACIP). MMWR Recomm Rep. Aug 11 2006;55(Rr-12):1-13.

14. Offit PA, Clark HF. RotaTeq: a pentavalent bovine--human reassortant rotavirus vaccine. Pediatr Ann. Jan 2006;35(1):29–34. doi:10.3928/0090-4481-20060101-11

15. Payne DC, Selvarangan R, Azimi PH, et al. Long-term Consistency in Rotavirus Vaccine Protection: RV5 and RV1 Vaccine Effectiveness in US Children, 2012–2013. Clinical Infectious Diseases. 2015;61(12):1792–1799. doi:10.1093/cid/civ872

16. Leshem E, Lopman B, Glass R, et al. Distribution of rotavirus strains and strain-specific effectiveness of the rotavirus vaccine after its introduction: a systematic review and meta-analysis. The Lancet Infectious Diseases. 2014/09/01/ 2014;14(9):847-856. 10.1016/S1473-3099(14)70832-1

17. Chua H, Feng S, Lewnard JA, et al. The Use of Test-negative Controls to Monitor Vaccine Effectiveness: A Systematic Review of Methodology. Epidemiology. 2020;31(1)

18. Payne DC, Staat MA, Edwards KM, et al. Active, Population-Based Surveillance for Severe Rotavirus Gastroenteritis in Children in the United States. Pediatrics. 2008;122(6):1235–1243. doi:10.1542/peds.2007-3378

19. Bowen MD, Mijatovic-Rustempasic S, Esona MD, et al. Rotavirus Strain Trends During the Postlicensure Vaccine Era: United States, 2008-2013. J Infect Dis. Sep 1 2016;214(5):732–8. doi:10.1093/infdis/jiw233

20. Stockdale JE, Liu P, Colijn C. The potential of genomics for infectious disease forecasting. Nature Microbiology. 2022/11/01 2022;7(11):1736-1743. doi:10.1038/s41564-022-01233-6

21. Organization WWH. Global genomic surveillance strategy for pathogens with pandemic and epidemic potential, 2022–2032. 2022;

22. Zeller M, Donato C, Trovão NS, et al. Genome-Wide Evolutionary Analyses of G1P[8] Strains Isolated Before and After Rotavirus Vaccine Introduction. Genome Biology and Evolution. 2015;7(9):2473–2483. doi:10.1093/gbe/evv157

23. Payne DC, Staat MA, Edwards KM, et al. Direct and Indirect Effects of Rotavirus Vaccination Upon Childhood Hospitalizations in 3 US Counties, 2006–2009. Clinical Infectious Diseases. 2011;53(3):245–253. doi:10.1093/cid/cir307

24. Zeller M, Heylen, E., Tamim, S., McAllen, J.K., Kirkness, E., Akopov, A., De Coster, S., Van Ranst, M., Matthijnssens, J. Data from: Comparative analysis of the Rotarix vaccine strain and G1P[8] rotaviruses isolated before and after vaccine introduction in Belgium. GenBank NCBI Virus. 2016. *Leuven, Belgium*.

25. Zeller M, Patton JT, Heylen E, et al. Genetic analyses reveal differences in the VP7 and VP4 antigenic epitopes between human rotaviruses circulating in Belgium and rotaviruses in Rotarix and RotaTeq. J Clin Microbiol. Mar 2012;50(3):966–76. doi:10.1128/jcm.05590-11

26. Matthijnssens J, Joelsson DB, Warakomski DJ, et al. Molecular and biological characterization of the 5 human-bovine rotavirus (WC3)-based reassortant strains of the pentavalent rotavirus vaccine, RotaTeq®. Virology. 2010/08/01/ 2010;403(2):111-127. 10.1016/j.virol.2010.04.004

27. Payne DC, Englund JA, Weinberg GA, et al. Association of Rotavirus Vaccination With Inpatient and Emergency Department Visits Among Children Seeking Care for Acute Gastroenteritis, 2010-2016. JAMA Network Open. 2019;2(9):e1912242-e1912242. doi:10.1001/jamanetworkopen.2019.12242

28. Simsek C, Bloemen M, Jansen D, et al. Rotavirus vaccine-derived cases in Belgium: Evidence for reversion of attenuating mutations and alternative causes of gastroenteritis. Vaccine. 2022/08/19/ 2022;40(35):5114-5125. 10.1016/j.vaccine.2022.06.082

29. Gilbert P, Self S, Rao M, Naficy A, Clemens J. Sieve analysis: methods for assessing from vaccine trial data how vaccine efficacy varies with genotypic and phenotypic pathogen variation. J Clin Epidemiol. Jan 2001;54(1):68–85. doi:10.1016/s0895-4356(00)00258-4

30. WHO. Guidance on conducting vaccine effectiveness evaluations in the setting of new SARS-CoV-2 variants: Interim guidance, 22 July 2021. Addendum to Evaluation of COVID-19 vaccine effectiveness. World Health Organization. 23 July 2021 2021;WHO REFERENCE NUMBER: WHO-2019-nCoV-vaccine-effectiveness-variants-2021.1.

31. Edlefsen PT, Rolland M, Hertz T, et al. Comprehensive Sieve Analysis of Breakthrough HIV-1 Sequences in the RV144 Vaccine Efficacy Trial. PLOS Computational Biology. 2015;11(2):e1003973. doi:10.1371/journal.pcbi.1003973

32. MacQueen JB. Some methods for classification and analysis of multivariate observations. presented at: Proceedings of the fifth Berkeley symposium on mathematical statistics and probability; 1968; California: University of California Press.

33. Sullivan SG, Tchetgen Tchetgen EJ, Cowling BJ. Theoretical Basis of the Test-Negative Study Design for Assessment of Influenza Vaccine Effectiveness. Am J Epidemiol. Sep 1 2016;184(5):345–53. doi:10.1093/aje/kww064

34. Cortese MM, Immergluck LC, Held M, et al. Effectiveness of monovalent and pentavalent rotavirus vaccine. Pediatrics. Jul 2013;132(1):e25–33. doi:10.1542/peds.2012-3804

35. Cates JE, Amin AB, Tate JE, Lopman B, Parashar U. Do Rotavirus Strains Affect Vaccine Effectiveness? A Systematic Review and Meta-analysis. Pediatr Infect Dis J. Dec 1 2021;40(12):1135–1143. doi:10.1097/inf.0000000000003286

36. Hoxie I, Dennehy JJ. Intragenic recombination influences rotavirus diversity and evolution. Virus Evolution. 2020;6(1)doi:10.1093/ve/vez059

37. Mhango C, Banda A, Chinyama E, et al. Comparative whole genome analysis reveals re-emergence of human Wa-like and DS-1-like G3 rotaviruses after Rotarix vaccine introduction in Malawi. Virus Evolution. 2023;9(1)doi:10.1093/ve/vead030

38. Hoxie I, Dennehy JJ. Rotavirus A Genome Segments Show Distinct Segregation and Codon Usage Patterns. Viruses. 2021;13(8):1460.

39. Duerr R, Dimartino D, Marier C, et al. Selective adaptation of SARS-CoV-2 Omicron under booster vaccine pressure: a multicentre observational study. eBioMedicine. 2023;97doi:10.1016/j.ebiom.2023.104843

40. Viboud C, Gostic K, Nelson MI, et al. Beyond clinical trials: Evolutionary and epidemiological considerations for development of a universal influenza vaccine. PLOS Pathogens. 2020;16(9):e1008583. doi:10.1371/journal.ppat.1008583

41. Pitzer VE, Bilcke J, Heylen E, et al. Did Large-Scale Vaccination Drive Changes in the Circulating Rotavirus Population in Belgium? Sci Rep. Dec 21 2015;5:18585. doi:10.1038/srep18585

42. Kirkwood CD, Boniface K, Barnes GL, Bishop RF. Distribution of Rotavirus Genotypes After Introduction of Rotavirus Vaccines, Rotarix® and RotaTeq®, into the National Immunization Program of Australia. The Pediatric Infectious Disease Journal. 2011;30(1):S48–S53. doi:10.1097/INF.0b013e3181fefd90

43. Hull JJ, Teel EN, Kerin TK, et al. United States rotavirus strain surveillance from 2005 to 2008: genotype prevalence before and after vaccine introduction. Pediatr Infect Dis J. Jan 2011;30(1 Suppl):S42–7. doi:10.1097/INF.0b013e3181fefd78

44. Roczo-Farkas S, Kirkwood CD, Cowley D, et al. The Impact of Rotavirus Vaccines on Genotype Diversity: A Comprehensive Analysis of 2 Decades of Australian Surveillance Data. J Infect Dis. Jul 13 2018;218(4):546–554. doi:10.1093/infdis/jiy197

45. Carvalho-Costa FA, Volotão EdM, de Assis RMS, et al. Laboratory-based Rotavirus Surveillance During the Introduction of a Vaccination Program, Brazil, 2005–2009. *The Pediatric Infectious Disease Journal*. 2011;30(1)

46. Matthijnssens J, Zeller M, Heylen E, et al. Higher proportion of G2P[4] rotaviruses in vaccinated hospitalized cases compared with unvaccinated hospitalized cases, despite high vaccine effectiveness against heterotypic G2P[4] rotaviruses. Clinical Microbiology and Infection. 2014/10/01/ 2014;20(10):O702-O710. 10.1111/1469-0691.12612

