## Supplemental Figure 1, Supplemental Figure 2, Supplemental Figures 3 & 4,Supplemental Table 1, Supplemental Figure 5, Supplemental Figure 6a for "Estimation of Rotavirus Vaccine Effectiveness Based on Whole Genome Sequences"

### Supplementary Methods

<u>Statistical model for sieve analysis</u>

The sieve analysis framework developed by Gilbert et al.[1] was modified to estimate the effect of vaccination status on the relative odds of infection with $\text{Strain}_{j=1,2,\ldots,K}$ versus $\text{Strain}_{j=0}$. As described in the main text, infected rotavirus strains were categorized into $K$ clusters using k-means clustering[2] across the distribution of genetic distance (GD) as defined by the percent nucleotide difference to the vaccine strain (or minimum distance to the five strains in the RotaTeq vaccine). The multinomial logistic regression model for sieve analysis for K strain categories[1] is given as follows:

$$\log\left\{\frac{Pr(J = j \mid clinically\ significant\ infection, V, x)}{Pr(J = 0 \mid clinically\ significant\ infection, V, x)}\right\} = \alpha_j + V\beta_j + x^T\omega_j$$

where $J \in \{0, 1, 2, \ldots, K\}$ denotes the infecting rotavirus strain, $V$ is the vaccination status ($V$=1, vaccinated; $V$= 0, unvaccinated) of the individual from whom strain $j$ was isolated, and $x$ denotes the vector of other individual-level covariates. For $j$=0, i.e. infection with a strain more closely related to the vaccine strain, we assume $\alpha_0 = \beta_0 = \omega_{0l} = 0$ for all $l$.

When only vaccination status among cases is considered (i.e., $V$ is the only covariate), parameter $\beta_j$ is the strain-specific log odds ratio (OR) and $\exp\{\beta_j\}$ represents the unadjusted OR ($J = j, J = 0$) for infection with $\text{Strain}_{j=1,2,\ldots,K}$ versus $\text{Strain}_{j=0}$ in a vaccinated individual compared to an unvaccinated individual.

### Supplemental Figures

#### Supplemental Figure 1.

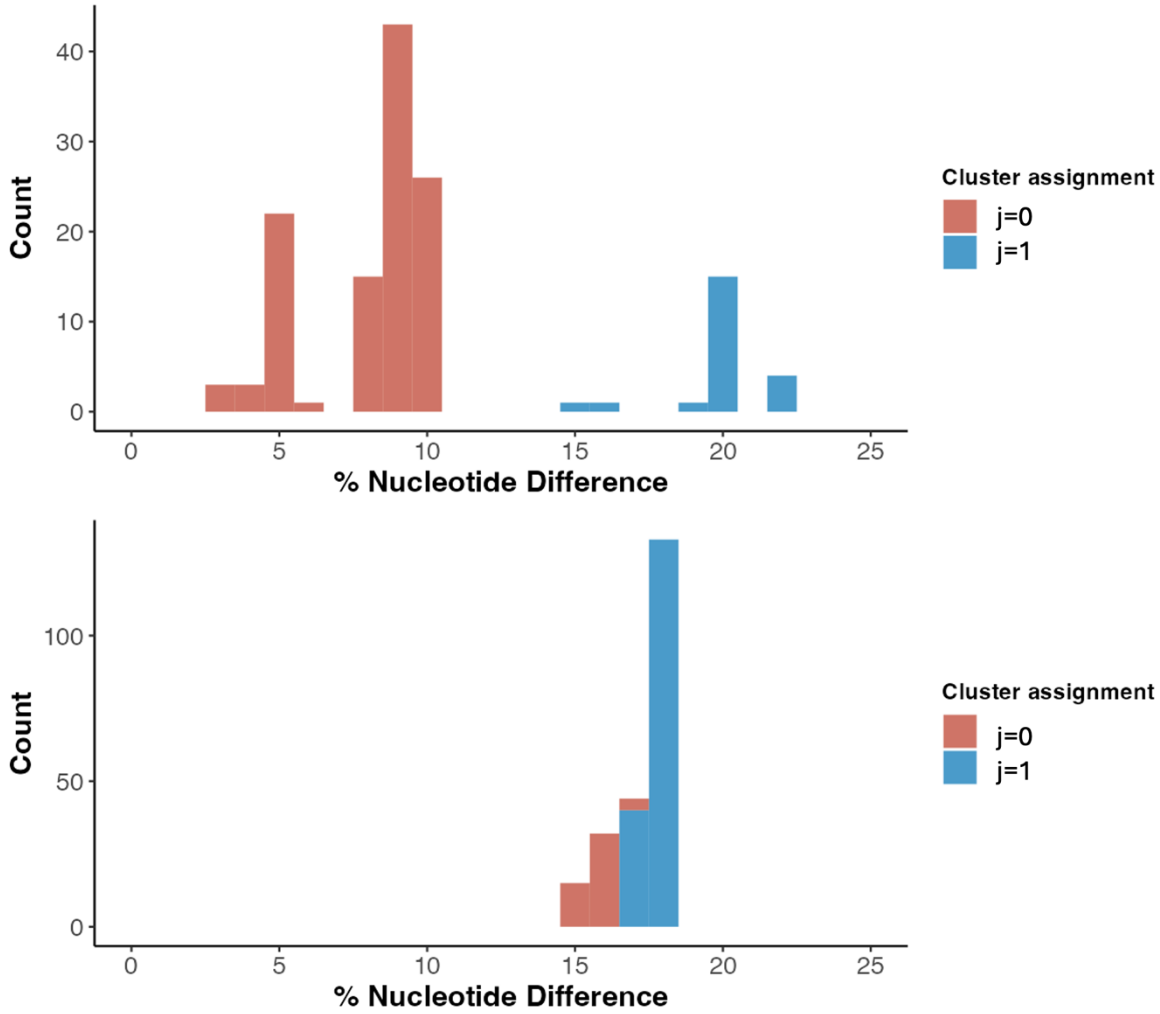

**Supplemental Figure 1. K-means clustering results.** Samples are colored by cluster assignment from the analysis. Includes samples in the (A) RV1-specific analysis and (B) RV5-specific analysis.

Supplemental Figure 2.

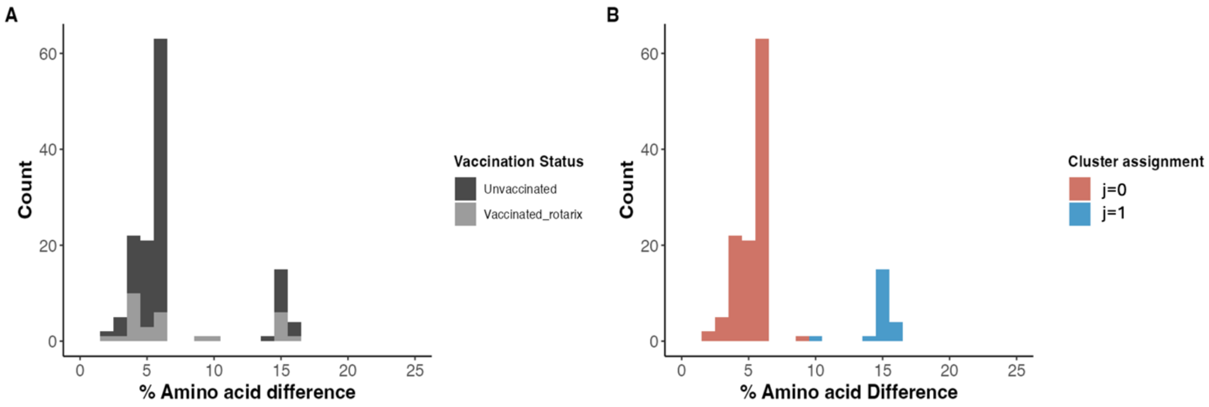

**Supplemental Figure 2. Sensitivity analysis using amino acid difference as genetic distance to RV1 vaccine strain, RV1-specific analysis.** The RV1-specific analysis includes cases who received at least one dose of RV1 vaccine or were unvaccinated. (A) Colored by vaccination status. (B) K-means clustering results. Samples are colored by cluster assignment from the analysis. The odds ratio of being infected with a strain above versus below the threshold for vaccinated versus unvaccinated individuals is 2.57 [95% confidence interval: 0.95, 6.97].

Supplemental Figure 3.

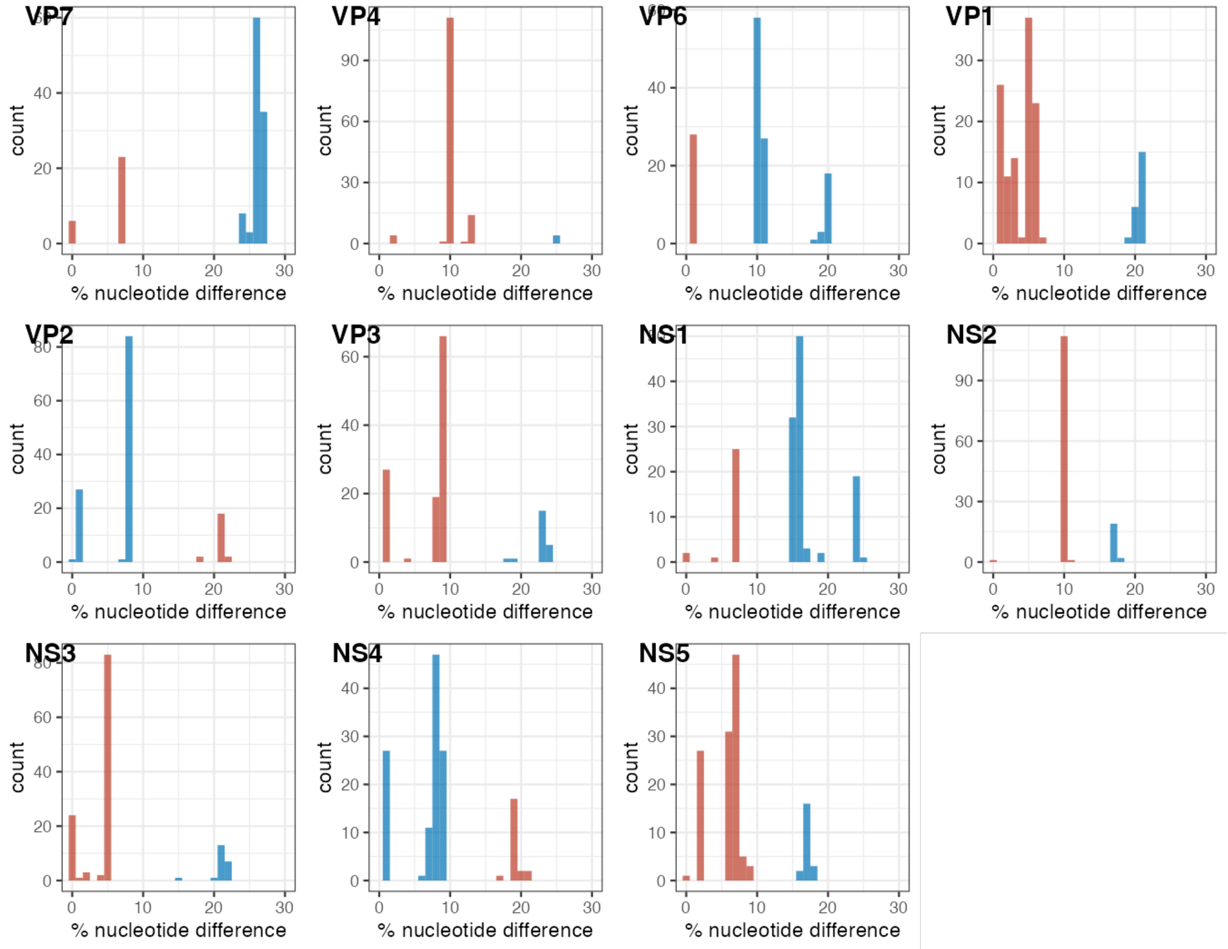

**Supplemental Figure 3. Individual segment-level K-means clustering results in the RV1-specific analysis.** Samples are colored by cluster assignment from the analysis. Cluster assignments are nominal, and the order of colors in this figure is arbitrary.

Supplemental Figure 4.

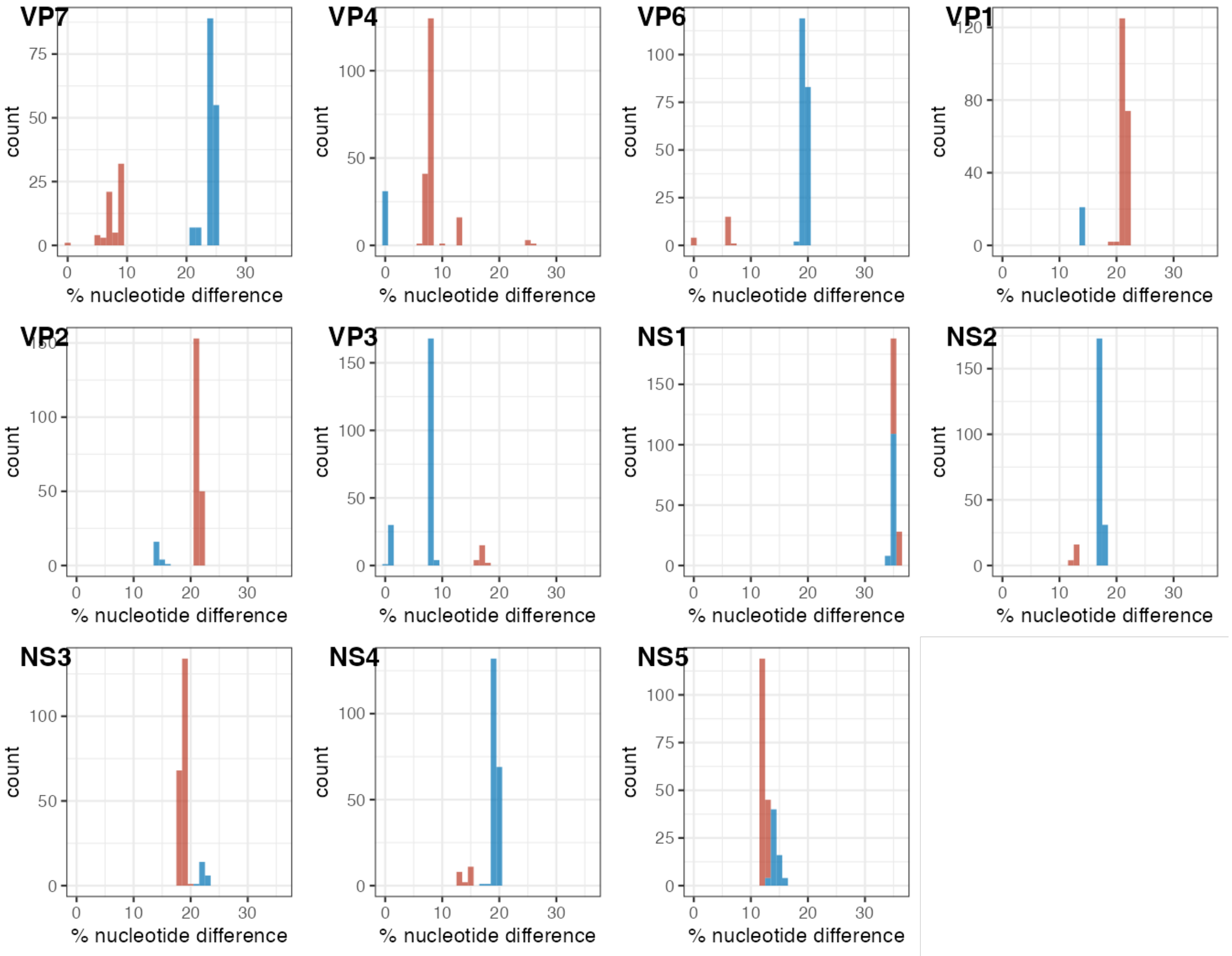

**Supplemental Figure 4. Individual segment-level K-means clustering results in the RV5-specific analysis.** Samples are colored by cluster assignment from the analysis. Cluster assignments are nominal, and the order of colors in this figure is arbitrary.

Supplemental Figure 5.

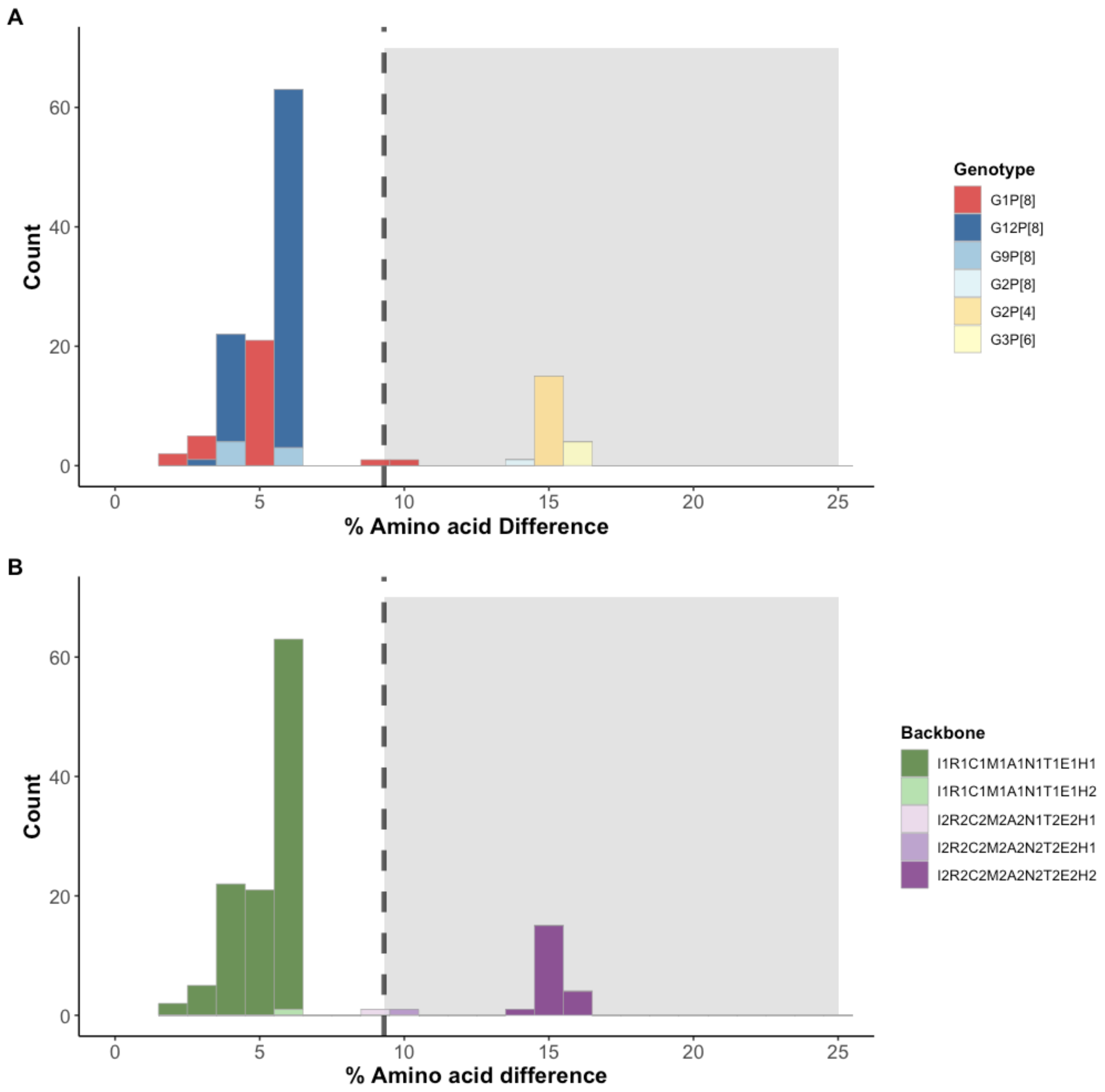

**Supplemental Figure 5. Characterization of the amino acid level genetic distance (GD) distribution for cases in the RV1-specific analysis according to genotype and genogroup.** Strains are colored according to (A) GxPx genotype information, and (B) genogroup of the remaining nine-segment backbone. GD is represented as percent amino acid difference between the case and RV1 vaccine strain. The black dotted line and grey background represent the threshold dividing $Strain_{j=0}$ vs $Strain_{j=1}$. The threshold is set at 9.3%.

Supplemental Figure 6.

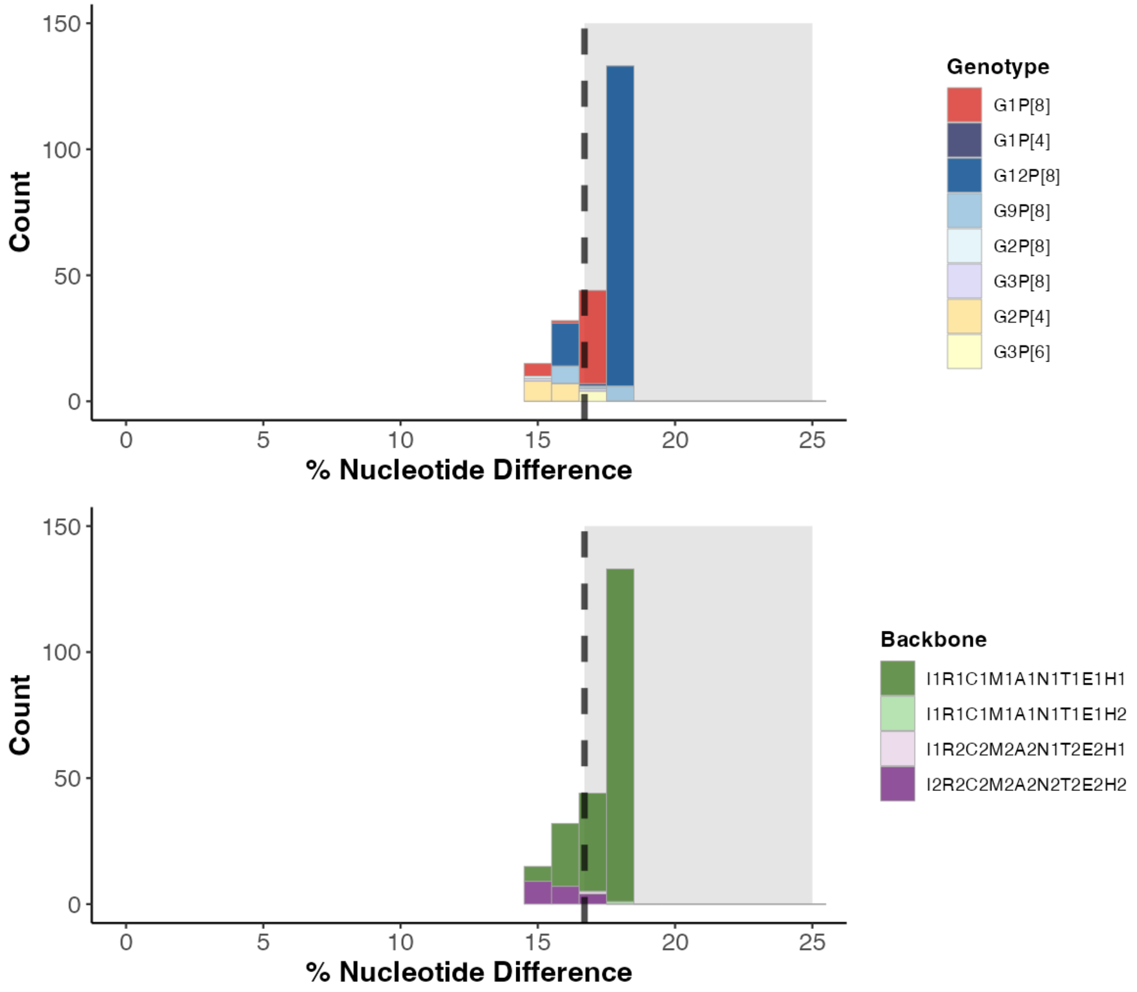

**Supplemental Figure 6. Characterization of the nucleotide-level genetic distance (GD) distribution for cases in the RV5-specific analysis according to genotype and genogroup.** Strains are colored according to (A) GxPx genotype information, and (B) genogroup of the remaining nine-segment backbone. GD is represented as minimum percent nucleotide difference between the case and to any of the five RV5 vaccine strains. The black dotted line and grey background represent the threshold dividing Strain $_{j=0}$ vs Strain $_{j=1}$. The threshold is set at 16.7%.

#### Supplemental Figure 7.

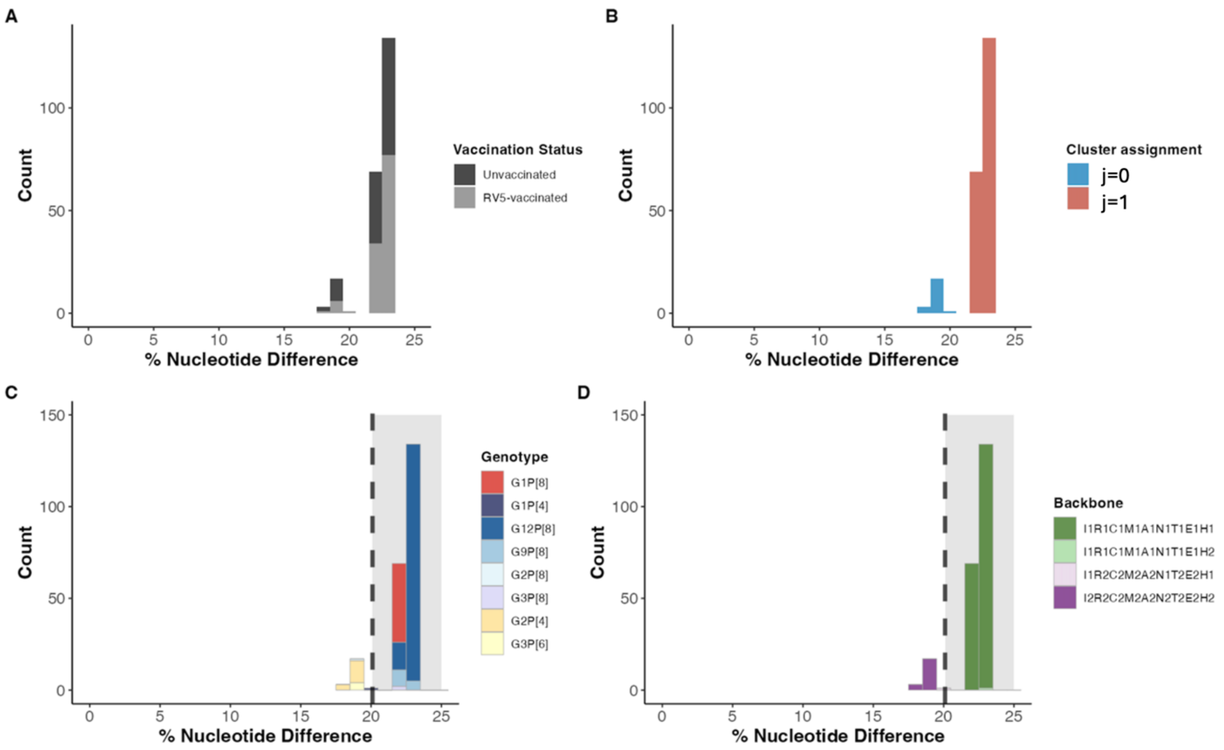

**Supplemental Figure 7. Sensitivity analysis using average distance to five RV5 strains, RV5-specific analysis.** Characterization of the nucleotide-level genetic distance (GD) distribution for cases in the RV5-specific analysis. GD is represented as average percent nucleotide difference between the case and to all of the five RV5 vaccine strains. Strains are colored according to (A) vaccination status, (B) cluster assignment in K-means clustering analysis, (C) GxPx genotype information, and (D) genogroup of the remaining nine-segment backbone. The black dotted line and grey background represent the threshold dividing Strain $_{j=0}$ vs Strain $_{j=1}$. The threshold is set at 20.1%.

Supplemental Figure 8

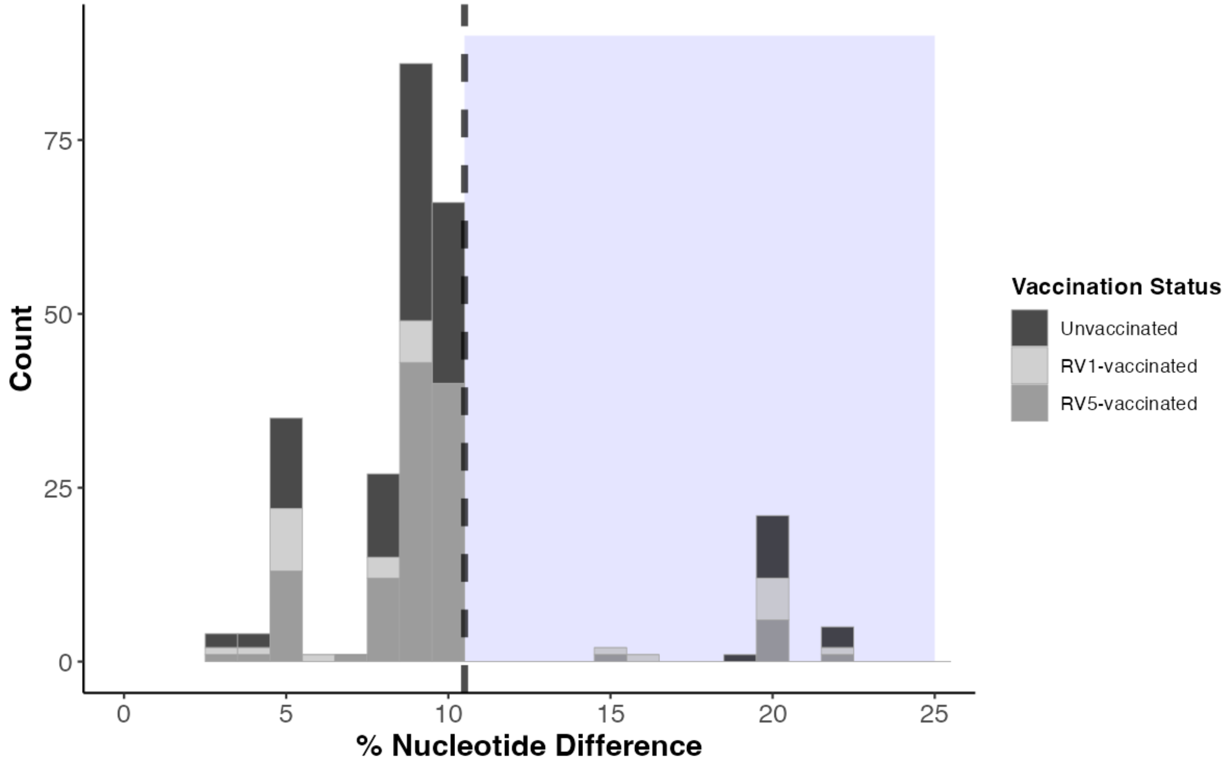

**Supplemental Figure 8. GD distribution of all RVGE cases to the RV1 vaccine strain colored by vaccination status.** All cases regardless of the vaccine type received – RV1-vaccinated, RV5-vaccinated, and Unvaccinated are included (n = 254). Genetic distance is represented as percent nucleotide difference between the case and RV1 vaccine strain. Black dotted line and blue background represents the threshold dividing between $Strain_{j=0}$ vs $Strain_{j=1.}$

Supplemental Figure 9.

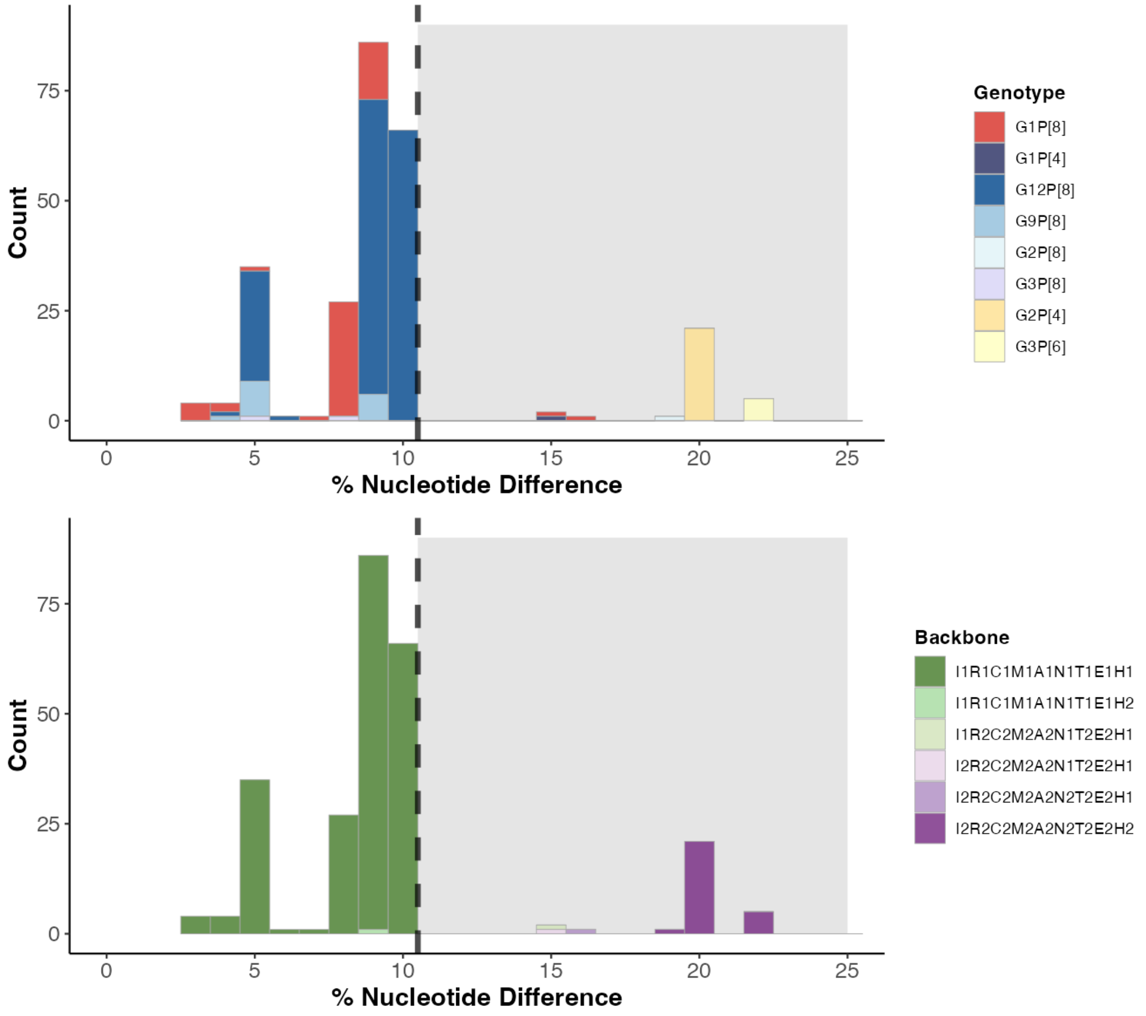

**Supplemental Figure 9. Nucleotide-level genetic distance (GD) distribution of all RVGE cases to the RV1 vaccine strain. Cases are colored by its (A) GxPx genotype information and (B) remaining nine segment backbone.** All cases regardless of the vaccine type received – RV1-vaccinated, RV5-vaccinated, and Unvaccinated are included (n = 254). GD is represented as percent nucleotide difference between the case and RV1 vaccine strain. Black dotted line and grey background represents the threshold dividing between $Strain_{j=0}$ vs $Strain_{j=1}$.

Supplemental Figure 10.

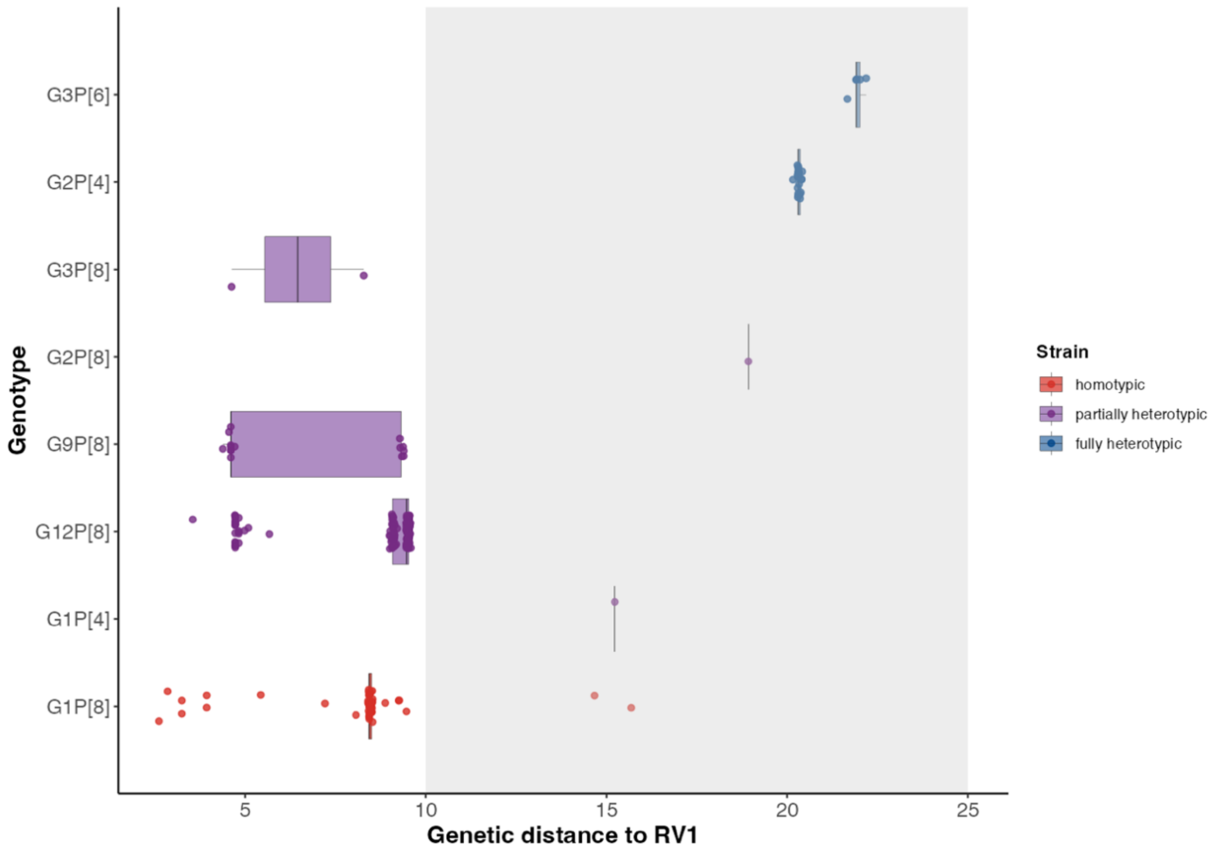

**Supplemental Figure 10. Genetic distance distribution for identified strains from each known GxPx genotype, overall analysis.** All cases regardless of the vaccine type received – RV1-vaccinated, RV5-vaccinated, and Unvaccinated are included. Genetic distance is represented as percent nucleotide difference between the case and RV1 vaccine strain. The grey background represents the threshold dividing $Strain_{j=0}$ vs $Strain_{j=1}$.

Supplemental Figure 11.

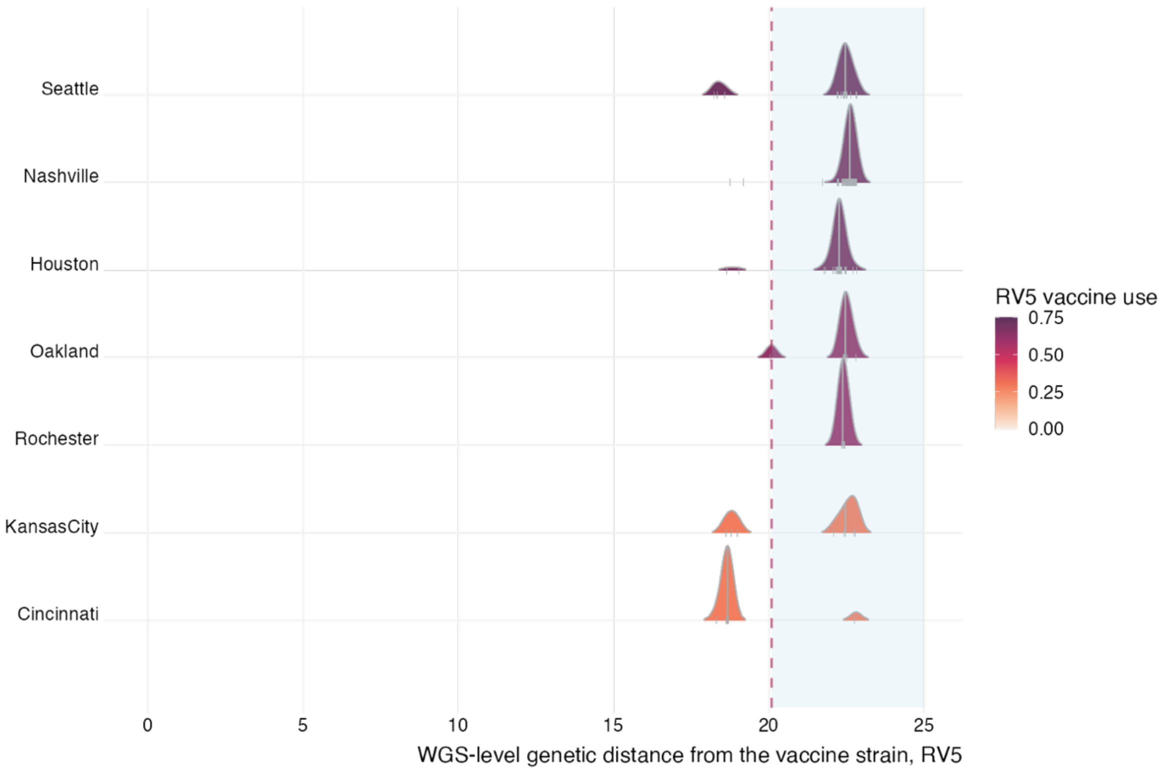

**Supplemental Figure 11. Average nucleotide-level genetic distance distribution of cases in RV5-specific analysis to all of RV5 vaccine strains for each of the seven U.S. NVSN study sites (n= 224).** Each site is colored by the proportion of vaccine usage amongst controls. The grey line in the distribution represents median GD for each study site. The red dotted line and light blue shaded area represents the threshold dividing $Strain_{j=0}$ vs $Strain_{j=1.}$ Study sites are ordered by descending vaccine use (top to bottom).

### Supplemental Tables

#### Supplemental Table 1.

**Supplemental Table 1. Individual segment-level sieve analysis results, vaccine-specific analysis.** Colors represent the direction of odds ratios for being infected with Strain$_{j=1}$ among vaccinated individuals. Segments colored in orange represent odds ratio (OR) > 1 and colored in blue represent OR < 1.

| Vaccine | Segment | Range | Threshold | Sieve analysis unadjusted OR [95% CI] | Segment length |
|---|---|---|---|---|---|
| RV1-specific analysis | VP7 | 0.31 – 27.42 | 7.5 | 1.06 [0.38, 2.91] | 981 |
| | VP4 | 1.71 - 24.95 | 12.9 | 1.17 [0.12, 11.70] | 2337 |
| | VP6 | 1.26 – 20.40 | 1.3 | 0.26 [0.11, 0.67] | 1191 |
| | VP1 | 0.92 - 21.18 | 7.2 | 3.03 [1.15, 8.02] | 3267 |
| | VP2 | 0.45 – 21.53 | 8.4 | 3.03 [1.15, 8.03] | 2694 |
| | VP3 | 1.47 – 24.07 | 8.9 | 3.03 [1.15, 8.03] | 2509 |
| | NSP1 | 0.00 – 24.73 | 7.5 | 0.26 [0.11, 0.67] | 1476 |
| | NSP2 | 0.00 – 18.13 | 10.6 | 2.57 [0.95, 6.97] | 954 |
| | NSP3 | 0.00 – 21.97 | 5.3 | 3.03 [1.15, 8.03] | 933 |
| | NSP4 | 1.14 – 21.02 | 9.5 | 3.03 [1.15, 8.03] | 528 |
| | NSP5 | 0.50 – 17.91 | 9.5 | 1.98 [0.72, 5.46] | 603 |
| RV5-specific analysis | VP7 | 0.00 – 24.87 | 8.9 | 1.54 [0.87, 2.76] | 981 |
| | VP4 | 0.00 – 25.72 | 0.1 | 1.45 [0.68, 3.10] | 2337 |
| | VP6 | 0.00 – 19.98 | 6.8 | 2.26 [0.87, 5.90] | 1191 |
| | VP1 | 13.53 – 22.07 | 14.4 | 1.96 [0.78, 4.93] | 3267 |
| | VP2 | 13.70 – 22.01 | 15.8 | 1.96 [0.78, 4.93] | 2694 |
| | VP3 | 0.08 – 17.90 | 8.6 | 0.51 [0.20, 1.28] | 2509 |
| | NSP1 | 33.54 – 35.70 | 35.0 | 1.31 [0.78, 2.22] | 1476 |
| | NSP2 | 12.26 – 18.13 | 13.3 | 2.26 [0.87, 5.90] | 954 |
| | NSP3 | 17.58 – 23.26 | 20.0 | 0.51 [0.20, 1.28] | 933 |
| | NSP4 | 12.50 – 20.08 | 15.4 | 1.96 [0.78, 4.94] | 528 |
| | NSP5 | 11.94 – 15.75 | 13.2 | 0.59 [0.33, 1.06] | 603 |

#### Supplemental Table 2.

**Supplemental Table 2. Genetic distance summary for each capsid-protein genotype (GxPx), RV1-specific analysis.** Genetic distance summary estimates are represented as percent nucleotide difference between the case and RV1 vaccine strain.

| **GxPx** | **n** | **Mean** (%) | **Median** (%) | **SD** (%) | **Minimum** (%) | **Maximum** (%) | **Relationship to RV1 strain** |
|---|---|---|---|---|---|---|---|
| G1P[8] | 29 | 7.97 | 8.44 | 2.80 | 2.86 | 15.68 | Homotypic |
| G12P[8] | 79 | 8.25 | 9.14 | 1.99 | 3.55 | 9.56 | Partially heterotypic |
| G9P[8] | 7 | 6.63 | 4.61 | 2.54 | 4.56 | 9.39 | Partially heterotypic |
| G2P[8] | 1 | 18.93 | 18.93 | NA | 18.93 | 18.93 | Partially heterotypic |
| G2P[4] | 15 | 20.32 | 20.31 | 0.07 | 20.16 | 20.42 | Fully heterotypic |
| G3P[6] | 4 | 21.87 | 21.91 | 0.15 | 21.66 | 22.01 | Fully heterotypic |

#### Supplemental Table 3.

**Supplemental Table 3. Vaccine effectiveness (VE) estimates against RVGE from study years 2012 – 2016, using binomial classification based on the two outer capsid proteins (GxPx).** In the RV1-specific analysis, only RV1-vaccinated and unvaccinated individuals were included. In the RV5-specific analysis, only RV5-vaccinated and unvaccinated individuals were included. Only GxPx genotypes with at least 10 cases were included in the VE estimation.

| **GxPx** | **RV1-specific analysis** | | **RV5-specific analysis** | |
|---|---|---|---|---|
| | Cases/controls | Adjusted* VE [95% CI] | Cases/controls | Adjusted* VE [95% CI] |
| G1P[8] | 29/5846 | 77.6% [42.3%, 92.3%] | 43/10125 | 70.31% [41.7%, 85.0%] |
| G2P[4] | 15/5846 | 64.0% [-14.8%, 89.0%] | 15/10125 | 84.0% [51.7%, 94.9%] |
| G9P[8] | 7/5846 | 91.0% [ 41.8%, 99.5%] | 14/10125 | 62.2% [-22.4%, 87.7%] |
| G12P[8] | 79/5846 | 75.0% [54.1%, 86.9%] | 144/10125 | 49.6% [25.2%, 65.8%] |

* Adjusted for age, care level (ED, inpatient, outpatient), collection year

#### Supplemental Table 4.

**Supplemental Table 4. Vaccine utilization breakdown among test-negative controls by study site.**

| | Controls (n = 12733) | | |
|---|---|---|---|
| **Study Site** | **Unvaccinated** (n = 3238) | **RV1-vaccinated** (n = 2608) | **RV5-vaccinated** (n = 6887) |
| Cincinnati | 489 (28) | 775 (44) | 492 (28) |
| Houston | 589 (27) | 116 (5) | 1513 (68) |
| Kansas City | 604 (26) | 1079 (46) | 663 (28) |
| Nashville | 730 (26) | 157 (6) | 1918 (68) |
| Oakland | 364 (23) | 253 (16) | 999 (62) |
| Rochester | 200 (21) | 170 (18) | 563 (60) |
| Seattle | 262 (25) | 58 (5) | 739 (70) |

#### Supplemental Table 5.

**Supplemental Table 5. Genetic distance-based strain distribution breakdown among RVGE cases by study site.**

| | **Genetic distance to the RV1 strain** Threshold: 9.6% (n = 254) | | **Genetic distance to the RV5 strain** Threshold: 16.7% (n = 254) | |
|---|---|---|---|---|
| **Study Site** | **Strain $_{j=0}$** (n = 224) | **Strain $_{j=1}$** (n = 30) | **Strain $_{j=0}$** (n= 70) | **Strain $_{j=1}$** (n= 184) |
| Cincinnati | 4 (21) | 15 (79) | 18 (95) | 1 (5) |
| Houston | 45 (96) | 2 (4) | 10 (21) | 37 (79) |
| Kansas City | 10 (67) | 5 (33) | 8 (53) | 7 (47) |
| Nashville | 134 (99) | 2 (1) | 8 (6) | 128 (94) |
| Oakland | 8 (89) | 1 (11) | 6 (67) | 3 (33) |
| Rochester | 10 (83) | 2 (17) | 8 (67) | 4 (33) |
| Seattle | 13 (81) | 3 (19) | 12 (75) | 4 (25) |

### References

1 Gilbert, P., Self, S., Rao, M., Naficy, A. & Clemens, J. Sieve analysis: methods for assessing from vaccine trial data how vaccine efficacy varies with genotypic and phenotypic pathogen variation. *J Clin Epidemiol* **54**, 68-85 (2001). https://doi.org/10.1016/s0895-4356(00)00258-4

2 MacQueen, J. B. in *Proceedings of the fifth Berkeley symposium on mathematical statistics and probability* Vol. (Vol. 1, pp. 281–297). (1968).
